# Education and Alzheimer Disease Genetic Risk in Associations of GLP-1 Receptor Agonists With Dementia Among Adults With Type 2 Diabetes

**DOI:** 10.64898/2026.09.04.26362304

**Authors:** Jingxuan Wang, Anna M. Pederson, Michael D. Flanders, Minhyuk Choi, Peter Buto, Kendra D. Sims, Ruijia Chen, Elyse Couch, Paola Gilsanz, Kaleen N. Hayes, Andrew R. Zullo, Andrew Stokes, M. Maria Glymour, Sarah F. Ackley

## Abstract

**Objective:** To evaluate whether educational attainment and Alzheimer disease genetic risk were associated with GLP-1 receptor agonist initiation and dementia incidence and whether adjustment for these measured factors materially changed the estimated association between GLP-1 receptor agonist initiation and incident dementia among adults with type 2 diabetes.

**Research Design and Methods:** We conducted an observational cohort study using linked electronic health record, survey, and genetic data from 14,364 All of Us Research Program participants with type 2 diabetes. We estimated associations of educational attainment and Alzheimer disease genetic risk with treatment initiation and incident dementia and compared GLP-1 receptor agonist initiators with initiators of non-sodium-glucose cotransporter 2 inhibitor second-line therapies, with a separate sodium-glucose cotransporter 2 inhibitor comparison. Models were estimated before and after additional adjustment for educational attainment, APOE ε4, and non-APOE genetic risk.

**Results:** Among 14,364 participants (mean age, 60.2 years; 54.2% female), the mean follow-up duration was 4.3 years. The estimated hazard ratio for dementia comparing GLP-1 receptor agonist initiation with non-SGLT2 inhibitor second-line therapy was 0.85 (95% CI 0.64-1.12) before adjustment for education or Alzheimer disease genetic risk and 0.84 (95% CI 0.64-1.12) after adjustment for educational attainment, APOE ε4, and non-APOE genetic risk.

**Conclusions:** Among adults with type 2 diabetes, adjustment for measured educational attainment and Alzheimer disease genetic susceptibility produced little change in the estimated association between GLP-1 receptor agonist initiation and incident dementia. These findings do not exclude confounding by these factors in other populations or residual confounding from socioeconomic, clinical, behavioral, and health-care-related factors.

**Highlights:**

- Observational studies have reported associations between GLP-1 receptor agonist use and lower dementia risk, but important social and genetic factors are often unavailable in electronic health record data.
- We examined whether educational attainment and Alzheimer disease genetic susceptibility were associated with treatment initiation and dementia and whether adjustment for these factors changed GLP-1 receptor agonist–dementia estimates.
- Adding measured education and genetic susceptibility produced little change in the estimated conditional hazard ratios.
- These results do not exclude residual confounding and should not be interpreted as evidence that GLP-1 receptor agonists prevent dementia.

## Introduction

Glucagon-like peptide-1 receptor agonists (GLP-1 RAs) and dual glucose-dependent insulinotropic polypeptide/GLP-1 receptor agonists (GIP/GLP-1 RAs) are widely used treatments for type 2 diabetes (T2D) and obesity.^1^ In addition to producing substantial weight loss,^2^ these agents improve glycemic control and reduce the incidence of hypertension, cardiovascular disease (CVD), liver and kidney disease, and other conditions associated with dementia risk.^3^ Preclinical and mechanistic studies further suggest that GLP-1 signaling may have direct neurobiological effects, including modulation of neuroinflammation.^4,5^ Together, these findings have motivated interest in the potential role of GLP-1-based therapies in dementia prevention.

Randomized controlled trial (RCT) evidence directly evaluating GLP-1-based therapies and dementia outcomes, however, remains limited. In the REWIND trial, there was some evidence dulaglutide was associated with reduced incident dementia risk; the intent-to-treat analysis estimated a protective point estimate (HR, 0.93; 95% CI, 0.85-1.02) though the effect was not statistically significant, and there was a statistically significant protective effect in a post-hoc covariate-adjusted analysis (HR, 0.86; 95% CI, 0.79-0.95), although this analysis was not prespecified.^6^ Trials designed to show effects of GLP-1RAs for cardiovascular outcomes also suggested a possible reduction in dementia risk (HR, 0.47; 95% CI, 0.25-0.86), although interpretation is limited by significant loss to followup and few total events (15 vs. 32).^7^ The ELAD trial did not show benefits of liraglutide treatment for its primary endpoint, cerebral glucose metabolism, despite post hoc analyses suggesting potential benefits on other neuroimaging outcomes.^8^ Most recently, the phase 3 evoke and evoke+ trials of oral semaglutide in 3800 adults with amyloid-positive mild cognitive impairment or early Alzheimer disease^9,10^ showed no statistically significant clinical benefit of treatment over placebo (HR for time to progression: (evoke, HR 0.98; 95% CI 0.85-1.14; evoke+, 0.96; 0.83-1.12).^11,12^ However, recent phase 3 trials evaluated treatment of biomarker-confirmed early symptomatic Alzheimer disease rather than dementia prevention among adults with type 2 diabetes, limiting direct comparison to observational studies.

Against this mixed randomized evidence, several observational studies using active-comparator, new-user designs have reported substantially more protective associations between GLP-1-based therapies and dementia risk, with estimates varying widely by comparator drug class: DPP4 inhibitors (HR, ∼0.58-0.83),^13–15^ SGLT2 inhibitors (SGLT2is, HR, ∼0.97-1.10),^16,17^ insulin (HR, ∼0.33),^18^ gliptins (HR, ∼0.52),^19^ and metformin (HR, ∼0.45).^20^ Because dementia is a relatively rare outcome and widespread use of GLP-1-based therapies is recent, these prior studies relied on large electronic health record (EHR) databases, such as TriNetX, to achieve sufficient statistical power.^20^ However, such databases often lack measures of key confounding variables.

The issue of unmeasured confounding is particularly relevant because use of GLP-1 and GLP-1/GIP RAs (*referred to as GLP-1 RAs henceforth)*, is strongly socially patterned, with higher uptake among higher socioeconomic status individuals.^21^ Inability to account for education may introduce residual socioeconomic confounding that contributes to discordance with trial findings. Education represents only one dimension of socioeconomic position. Health care access and utilization, income and wealth, insurance coverage, neighborhood context, social stressors and supports, alcohol use, and detailed cardiometabolic characteristics may also influence treatment selection and dementia risk. However, higher educational attainment is a well-established and strong predictor of reduced dementia risk and predicts better than other socioeconomic factors.^22^

In addition, genetic susceptibility to Alzheimer disease (AD), a common form of dementia, may confound associations between GLP-1 RA initiation and dementia, but is typically unmeasured in large EHR databases. Genetic risk for AD is associated with midlife declines in body mass index (BMI),^23–25^ lower blood pressure in later life^26^ and lower blood glucose,^25^ which may reduce the risk of GLP-1 RA initiation. Conversely, carriers of the apolipoprotein E ε4 (*APOE*-ε4) allele—the strongest known genetic risk factor for AD—have higher levels of low-density lipoprotein cholesterol (LDL-C) and other cardiovascular risk factors that may affect the probability GLP-1 RA initiation.^25^ Together, these pathways raise concern that observed associations may not reflect true neuroprotective effects.

Whether the substantially more favorable associations observed in large observational studies reflect true neuroprotective effects or residual confounding therefore remains uncertain. Understanding potential sources of differences across studies is important for interpreting the rapidly evolving evidence on GLP-1-based therapies and dementia. In addition, clarifying this discrepancy is critical for interpreting emerging data and for patients considering GLP-1-based therapies for the prevention of dementia. Cognitive impairment and dementia can complicate diabetes self-management, medication adherence, and safe glycemic treatment. Observational claims of cognitive benefit may also influence selection of glucose-lowering therapies, making the validity and interpretation of these associations clinically relevant for diabetes care.

In this study, we used the All of Us Research Program, which integrates electronic health record, survey, and genetic data, to evaluate potential confounding by factors usually unavailable in large electronic health record databases. Among adults with type 2 diabetes, we examined whether (1) educational attainment and Alzheimer disease genetic risk were associated with GLP-1 receptor agonist initiation, (2) these factors were associated with incident dementia, and (3) adjustment for these factors changed the estimated association between GLP-1 receptor agonist initiation and incident dementia.^27^ The present study focuses specifically on use of these therapies among adults with type 2 diabetes and therefore does not address GLP-1 receptor agonist use solely for obesity.

## Research Design and Methods

### Overview

We first evaluated whether education and AD genetic risk were associated with GLP-1 RA initiation among All of Us participants with T2D. Second, we evaluated whether dementia incidence was associated with education and AD genetic risk. Third, we used an active-comparator, new-user (ACNU) design^28,29^ to compare initiation of GLP-1 RAs versus other second-line glucose-lowering therapies in relation to incident dementia, with follow-up aligned at treatment initiation. An ACNU design compares patients initiating a treatment to patients initiating a clinically relevant alternative, aligning follow-up at treatment start to reduce confounding and time-related biases.^30^ Associations were estimated sequentially with and without adjustment for education and Alzheimer disease genetic risk to quantify changes in the estimated treatment-outcome association after incorporating these measured factors.

### Study Sample

The All of Us research program^31^ began recruitment of US adults in May of 2018 at more than 340 recruitment sites.^31^ Although All of Us recruitment began in May 2018, linked electronic health record data included retrospectively collected clinical information predating enrollment. Approximately 95% of All of Us participants have consented to share existing and retrospectively collected EHR data, which allows researchers to follow the majority of participants for health outcomes such as dementia. EHR data across sites is harmonized using the Observational Medical Outcomes Partnership (OMOP) Common Data Model.^32^ All of Us obtains informed consent from all participants and provides documentation of consent procedures. Because this study used deidentified data, it was not considered human subjects research. The present data management and analyses were exempt from institutional review board review because we used deidentified data. We followed the Strengthening the Reporting of Observational Studies in Epidemiology (STROBE) reporting guideline for cohort studies.

### Inclusion and Exclusion

Analyses were restricted to participants with linked electronic health record and genetic data, nonmissing educational attainment, and an electronic health record-recorded type 2 diabetes diagnosis between January 1981 and September 2023 (definition in **eTable 1**). We included participants with prescription of GLP-1/GIP RAs or other second-line diabetes treatments. We excluded individuals with GLP-1 RA or second-line drug initiation prior to a T2D diagnosis. Individuals with pancreatitis, thyroid cancer, type 1 diabetes, gastroparesis, AD medication use, all-cause dementia, or traumatic brain injury prior to initiation of GLP-1/GIP RA or comparator drug were also excluded. Details regarding diagnostic codes and Observational Medical Outcomes Partnership (OMOP) concept names used to implement inclusions and exclusions are provided in **eTable 1**. Flowchart of inclusion and exclusion is provided in **eFigure1**.

### Treatment Groups

We compared individuals initiating GLP-1 RAs with two comparator groups: those initiating SGLT2is and those initiating any other second-line treatments for T2D. Other second-line treatments include the following drug classes: DPP4 inhibitors (gliptans), sulfonylureas, meglitinides, thiazolidinediones, alpha-glucosidase inhibitors, an amylin analog, a bile acid sequestrant, and a dopamine agonist. These therapies were grouped to represent non-SGLT2 inhibitor second-line glucose-lowering treatment. We separated out SGLT2is due to their beneficial effects on cardiovascular risk and BMI.^33^ Also consistent with prior studies,^19^ we controlled for history of metformin or insulin use. More details are provided in **eTable 1**. Participants initiating a comparator therapy who subsequently initiated a GLP-1 receptor agonist contributed to the comparator group until GLP-1 receptor agonist initiation and were classified in the GLP-1 receptor agonist group thereafter.

### Definition of Index Date and Treatment and Comparator Groups

Medication exposure was based on electronic health record prescription records; these data indicate prescribing rather than confirmed dispensing, adherence, or medication use. The index date was defined as the date of GLP-1 RA or second-line diabetes drug initiation. Individuals were followed from the index date to the earliest date of dementia diagnosis, death, or censoring. Additional details on lookback periods and prescription data are given in the **Supplemental Methods**.

### AD Genetic Risk

AD genetic risk was operationalized in two ways: number of *APOE*-ε4 alleles and AD genetic risk score, capturing non-APOE-ε4 genetic risk. *APOE* haplotype was derived using the single nucleotide polymorphisms (SNPs) rs7412 and rs429358. The AD genetic risk score was constructed using the lead SNPs from 83 non-*APOE*-ε4 loci present in All of Us that reached genome-wide significance in the most recent meta-analysis of AD genetic risk.^34^ The AD genetic risk score was calculated by taking the sum of the product of risk allele counts and GWAS-derived coefficients for log-odds of AD. Additional details on the construction and validation of this AD genetic risk score are described elsewhere.^35^

### Dementia Ascertainment

Dementia outcomes included Alzheimer disease, vascular dementia, Lewy body dementia, other specified dementias, and unspecified dementia, defined using International Classification of Diseases, Ninth and Tenth Revision (ICD-9 and ICD-10) codes in the EHR, along with corresponding Systematized Nomenclature of Medicine (SNOMED) codes. When multiple diagnosis dates were recorded, we used the earliest date. A complete list of codes is provided in **eTable 1**. Justification for why dementia subtypes were merged is given in the appendix.

### Covariates and Confounders

Models examining associations between education and GLP-1 RA initiation adjust for age, quadratic age (age squared), self-reported sex-assigned at birth, continuous BMI (at the time of the survey), as well as self-reported categorical race and ethnicity. Models included age and age squared at treatment initiation to allow a nonlinear association between age and the outcomes.

Race and ethnicity were categorized as non-Hispanic White, non-Hispanic Black, non-Hispanic Asian, Hispanic, and other or missing. Models examining associations between AD genetic risk and GLP-1 RA initiation additionally adjusted genetic ancestry operationalized as the first 5 genetic principal components. Educational attainment was obtained from participant survey responses and categorized as less than high school, high school, some college, or college or above to avoid imposing a linear relationship across educational credentials.

Models estimating the association between GLP-1 RA initiation and incident dementia are adjusted for age and quadratic at drug initiation, sex, as well as race and ethnicity as assessed. Additionally, models account for the presence of comorbidities, including obesity-related diagnoses, CVD, depression, diabetes complications, dyslipidemia, hypertension, liver disease, sensory impairment, sleep disorders, tobacco use, and the use of insulins or metformin. Comorbidities were identified from electronic health record diagnoses using code definitions listed in **eTable 1**. The primary treatment-outcome models included obesity-related diagnoses rather than continuous BMI to account for BMI at the time of drug initiation, which may have preceded the survey. A full list of confounders is given in **eTable 1**.

### Statistical Analysis

First, we estimated the associations of education and AD genetic risk with GLP-1 RA initiation in separate models. Modified-Poisson regression models estimated the risk of use by categorical education level, *APOE*-ε4 allele count, and AD genetic risk score.^36^ Second, to characterize potential confounding pathways, we examined whether education and AD genetic risk were associated with dementia incidence. Separate Cox proportional-hazards models were used to estimate the association of education and AD genetic risk with dementia incidence (details in **Supplemental Methods**). Consistent with prior studies,^13,14,19^ we used Cox proportional-hazards models. The base model adjusted for only covariates listed in **eTable 1**. Subsequent models adjusted for education, *APOE*-ε4, and non-APOE-ε4 AD genetic risk, and combinations therein. We calculated ratios of hazard ratios (relative hazard ratios, RHR) to describe the relative change in conditional hazard ratio estimates after additional adjustment for education and Alzheimer disease genetic risk. Because hazard ratios are non-collapsible, these ratios were treated as descriptive comparisons rather than formal measures of confounding. Bootstrapping with 1000 replicates was used to assess statistical uncertainty. We conducted all analyses using R. Statistical analyses were conducted from October 22, 2024 to June 5, 2026.

### Stability Analyses

We conducted four stability analyses. First, because younger individuals are unlikely to be at risk for dementia, we restricted the analysis to individuals aged 55 years or older at treatment initiation. Second, because EHR data from All of Us may be incomplete (EHR data from All of Us may have gaps in longitudinal capture when individuals receive care outside participating health systems),^37^ we restricted the analysis to a subcohort of individuals at medical centers with more consistent follow-up (**Supplemental Methods**). Third, because obesity may be a confounder, we adjusted for obesity within two years before treatment initiation. Finally, to more closely align the study population with the evoke and evoke+ trials,^12^ which enrolled participants with mild cognitive impairment or mild dementia, we restricted the analysis to individuals with a mild cognitive impairment diagnosis before treatment initiation.

### Data and Resource Availability

Data are available to authorized researchers through the All of Us Researcher Workbench, subject to program access requirements.

## Results

At the time of analysis (March 2026), 393,600 All-of-Us participants had EHR data available. Of these, 14,364 had a T2D diagnosis before initiation of any of GLP-1 RA, SGLT2i, or other comparator second-line drugs and met all other inclusion criteria. Among these participants, the mean baseline age was 60.2 years, 54.2% were female, 30.9% had some college education, and 33.0% had a college degree or higher. In the ACNU analysis with the same 14,364 participants, 6,817 participants initiated a GLP-1, 2,547 initiated an SGLT2i, and 5,000 initiated other comparator second-line drugs. During the mean follow up of 4.3 years, 364 incident dementia cases occurred. **Table 1** gives the demographic characteristics of these two cohorts. GLP-1 RA initiators are younger, more likely to be female, and less likely to have cardiovascular and cerebrovascular disease.

**Table 1:**
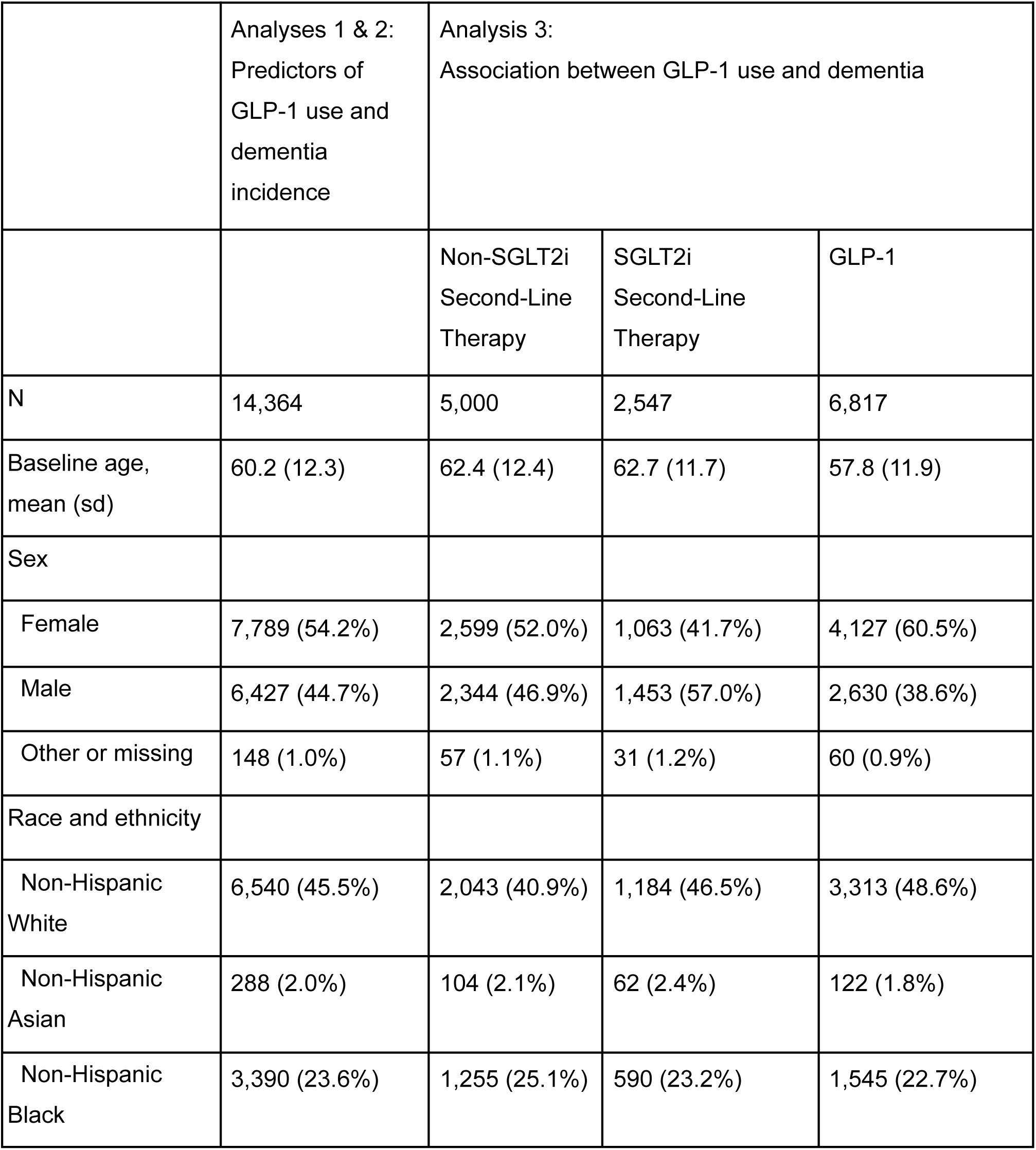
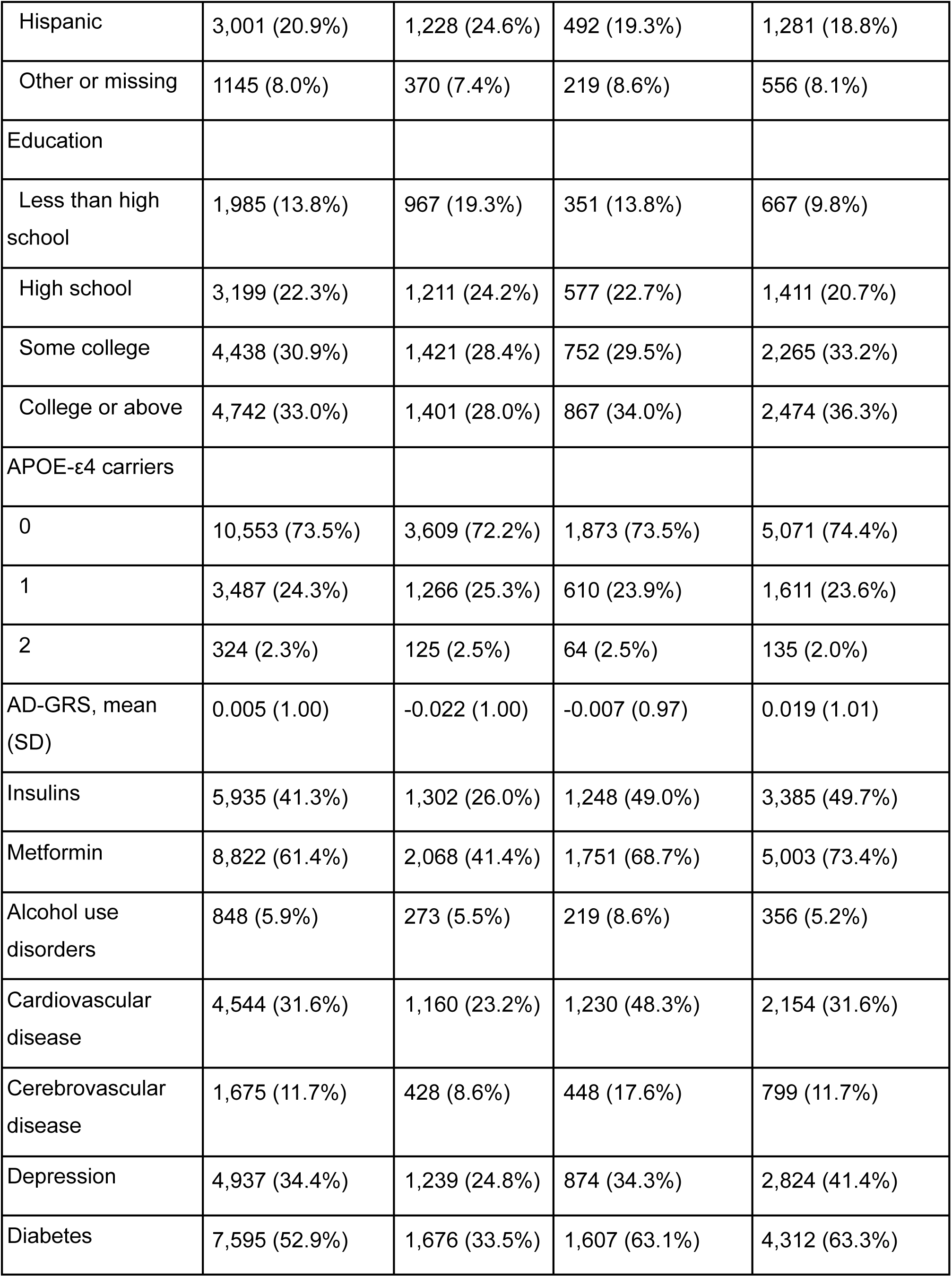
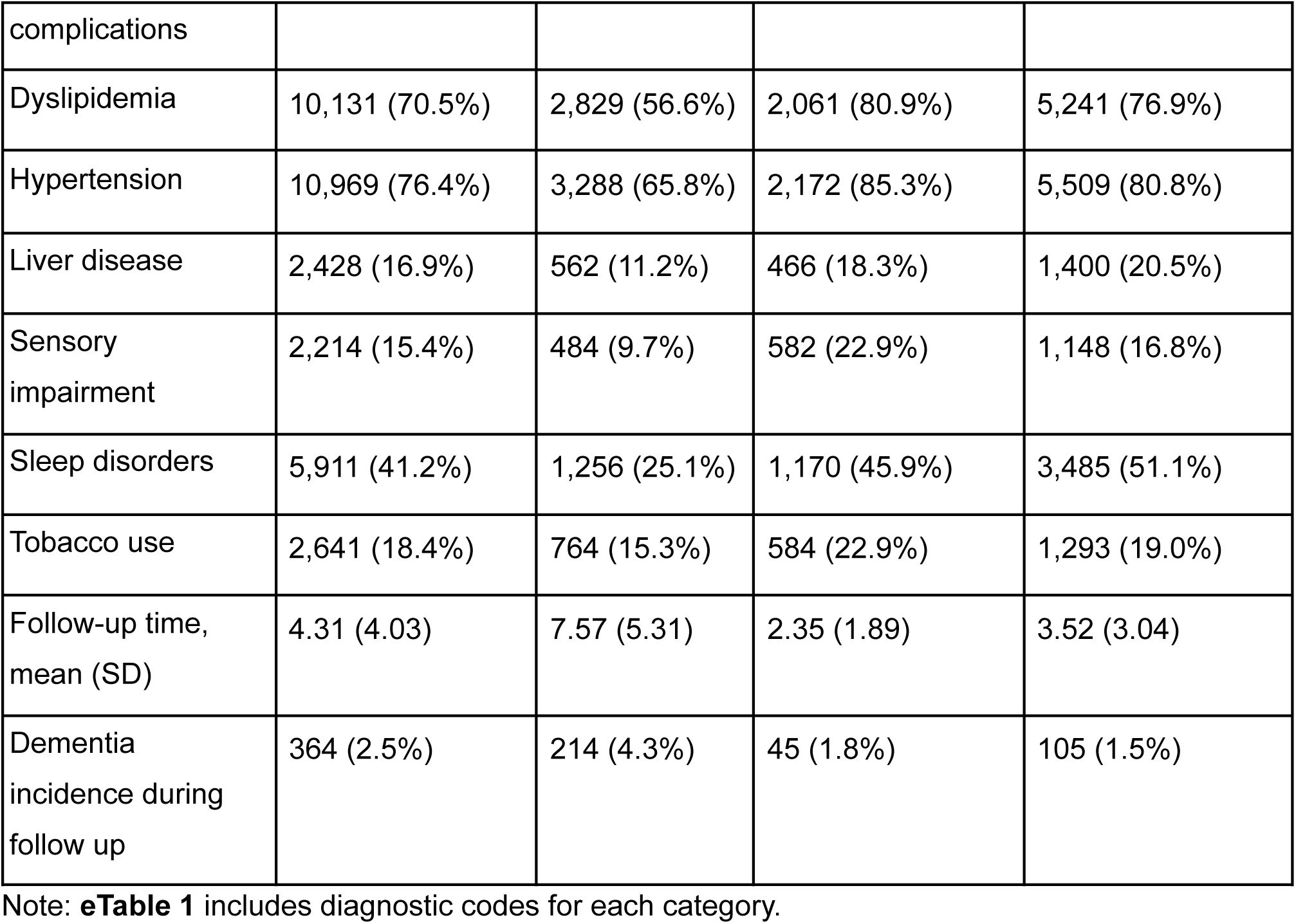
Characteristics of the overall analytic cohort and participants at initiation of non-SGLT2 inhibitor, SGLT2 inhibitor, or GLP-1 receptor agonist therapy.

### Associations of Education and Genetics with GLP-1 RA Initiation

Compared to individuals with less than high school education, GLP-1 RA initiation over a non-SGLT2i or a SGLT2i second-line therapy was positively associated with having completed high school (Figure 2, RR GLP-1 Initiation = 1.23, 95% CI: 1.14-1.33), some college (RR GLP-1 Initiation = 1.41, 95% CI: 1.31-1.51), and college or higher education (RR GLP-1 Initiation = 1.53, 95% CI: 1.43-1.65). Compared to non-*APOE*-ε4 carriers, one and two *APOE*-ε4 alleles were associated with a lower likelihood of GLP-1 RA initiation (RR GLP-1 Initiation = 0.94, 95% CI: 0.90-0.98 and RR GLP-1 Initiation = 0.84, 95% CI: 0.74-0.95, respectively). A one standard deviation increase in non-*APOE*-ε4 AD genetic risk was not associated with the risk of GLP-1 RA initiation (RR GLP-1 Initiation = 1.00, 95% CI: 0.98-1.02). These results are summarized in **Figure 2**.

**Figure 1:**
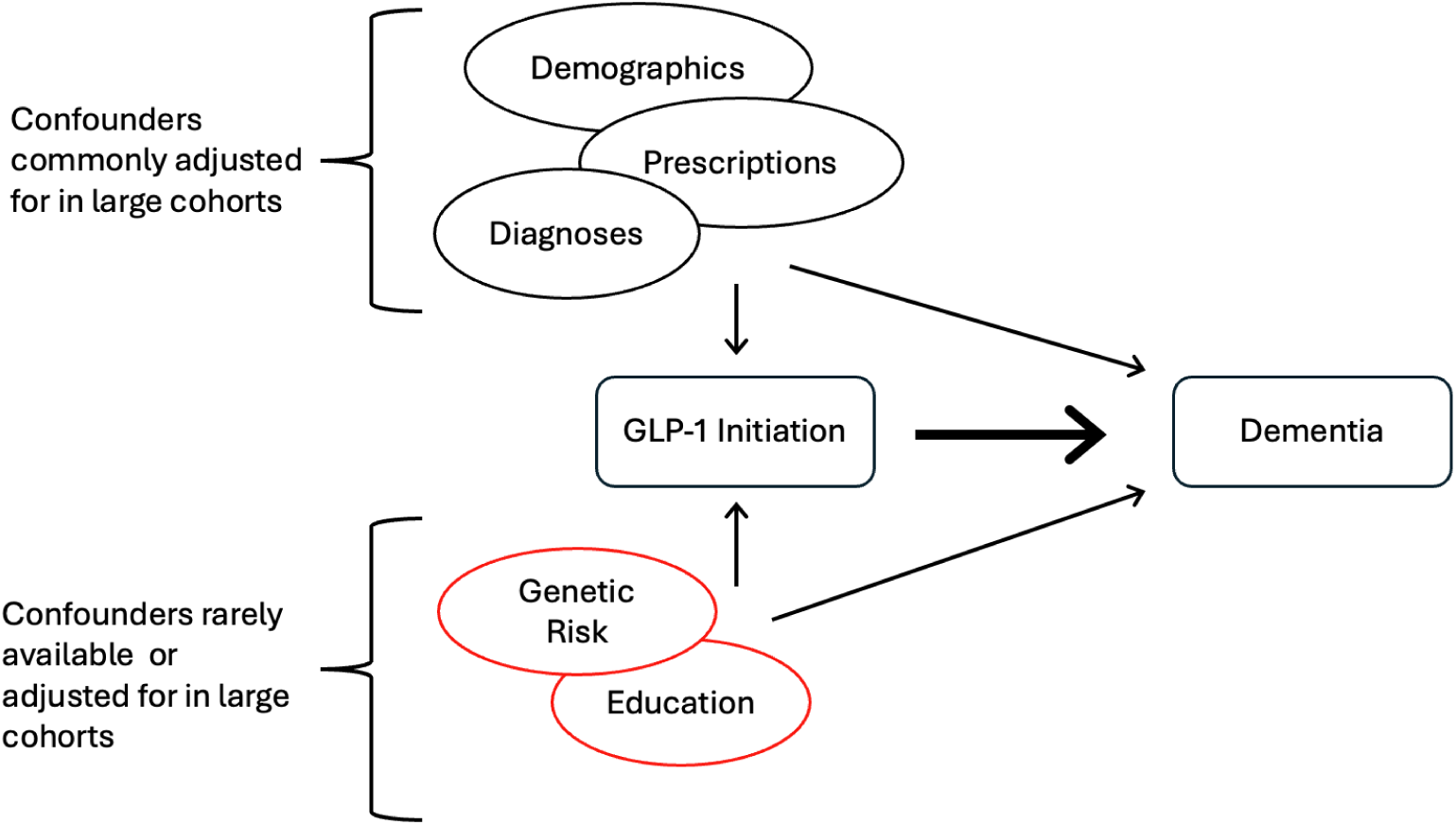
Conceptual model of the study. We evaluated educational attainment and Alzheimer disease genetic susceptibility as potential confounders of associations between GLP-1 receptor agonist initiation and dementia. These factors are often unavailable in electronic health record-based studies; additional socioeconomic, clinical, health care access, and behavioral factors may also contribute to confounding.

**Figure 2.**
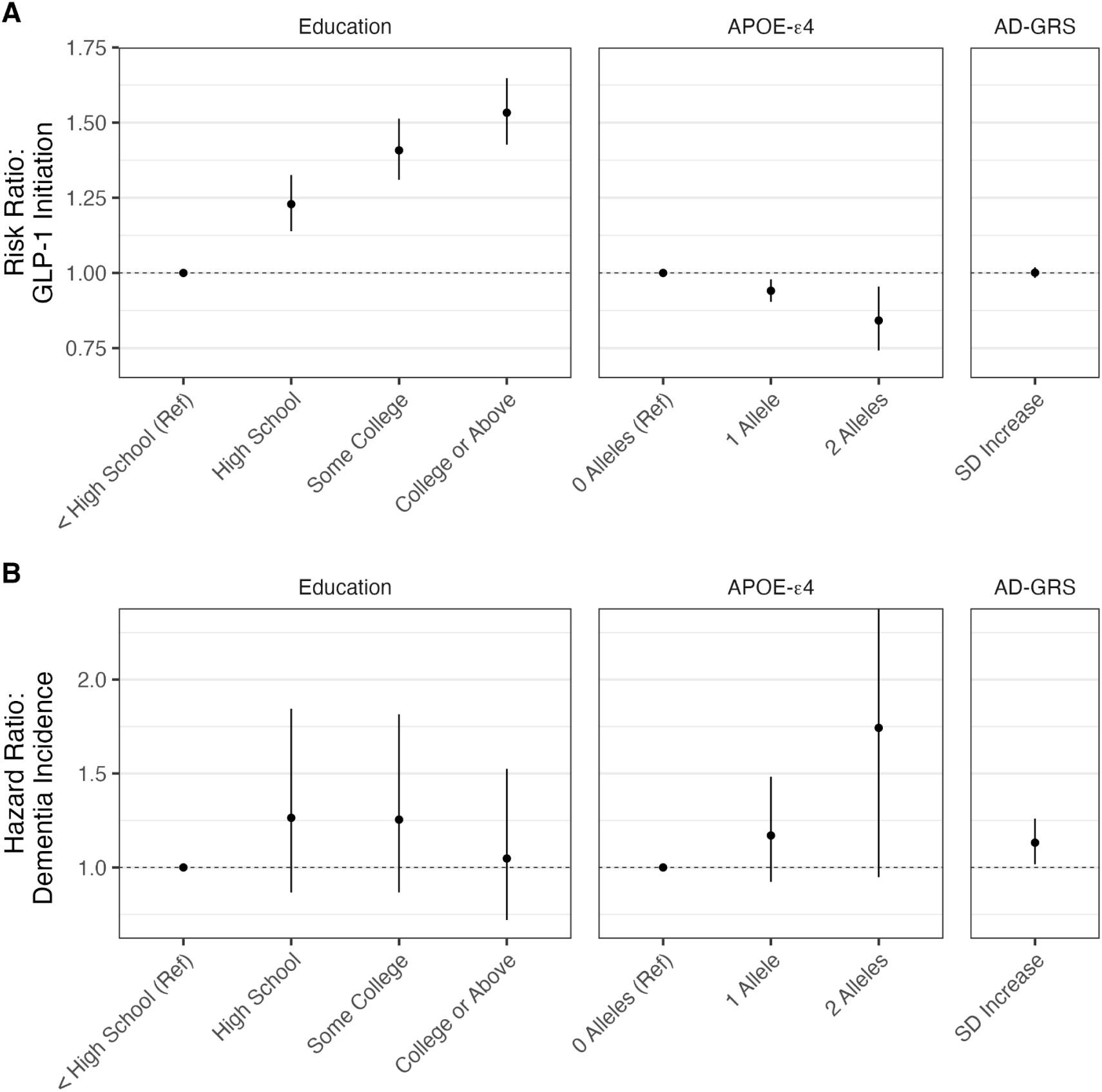
Typically unmeasured variables that are potential confounders for GLP-1 RA initiation and dementia incidence. (A) Relative risk of initiation of GLP-1 receptor agonists for education, APOE-ε4 genotype, and AD genetic risk score (AD-GRS). (B) Relative hazards for dementia incidence for the same variables. Points indicate point estimates and vertical lines indicate 95% confidence intervals. Reference categories are shown at risk or hazard ratio equal to 1.

### Associations of Education and Genetics with Time-to-Dementia

Educational attainment was not associated with dementia incidence (**Figure 2**). Compared to individuals with less than high school education, HRs were 1.26 (95% CI: 0.87-1.84) for high school, 1.25 (95% CI: 0.87-1.82) for some college, and 1.05 (95% CI: 0.72-1.53) for college or higher education. Point estimates for dementia hazard were higher among participants with one APOE ε4 allele (HR 1.17, 95% CI 0.92-1.48) and two alleles (HR 1.74, 95% CI 0.95-3.20), although the 95% confidence intervals included the null. A one standard deviation increase in non-*APOE* AD genetic risk score was associated with a modest increase in dementia hazard (HR = 1.13, 95% CI: 1.02-1.26).

### Associations of GLP-1 RA Initiation with Time-to-Dementia

In the model unadjusted for education or AD genetic risk but adjusted for a standard set of EHR-available confounders (see Methods), GLP-1 RA initiation compared with a non-SGLT2i second-line therapy was associated with reduced dementia incidence (HR = 0.85, 95% CI: 0.64-1.12) (**Figure 3**), but this reduction was not statistically significant. Similar associations adjusted for a standard set of EHR-available confounders were observed when comparing GLP-1 RA with SGLT2i initiation (HR =0.90 95% CI: 0.61-1.33) (**eFigure 2**). Adjustment for education and AD genetic risk did not change the estimated association of GLP-1 RA initiation on dementia incidence (**Figures 3**). For example, adjustment for educational attainment, APOE-ε4, non-APOE genetic risk, or combinations of these factors minimally changed the estimates of GLP-1 RA initiation compared with a non-SGLT2i second-line therapy (fully adjusted HR=0.84; 95% CI, 0.64-1.12). Point estimates from stability analyses were generally directionally consistent with the primary analysis, although the estimate was more protective in the restriction to sites with more consistent electronic health record follow-up (**eFigures 3-10**).

**Figure 3.**
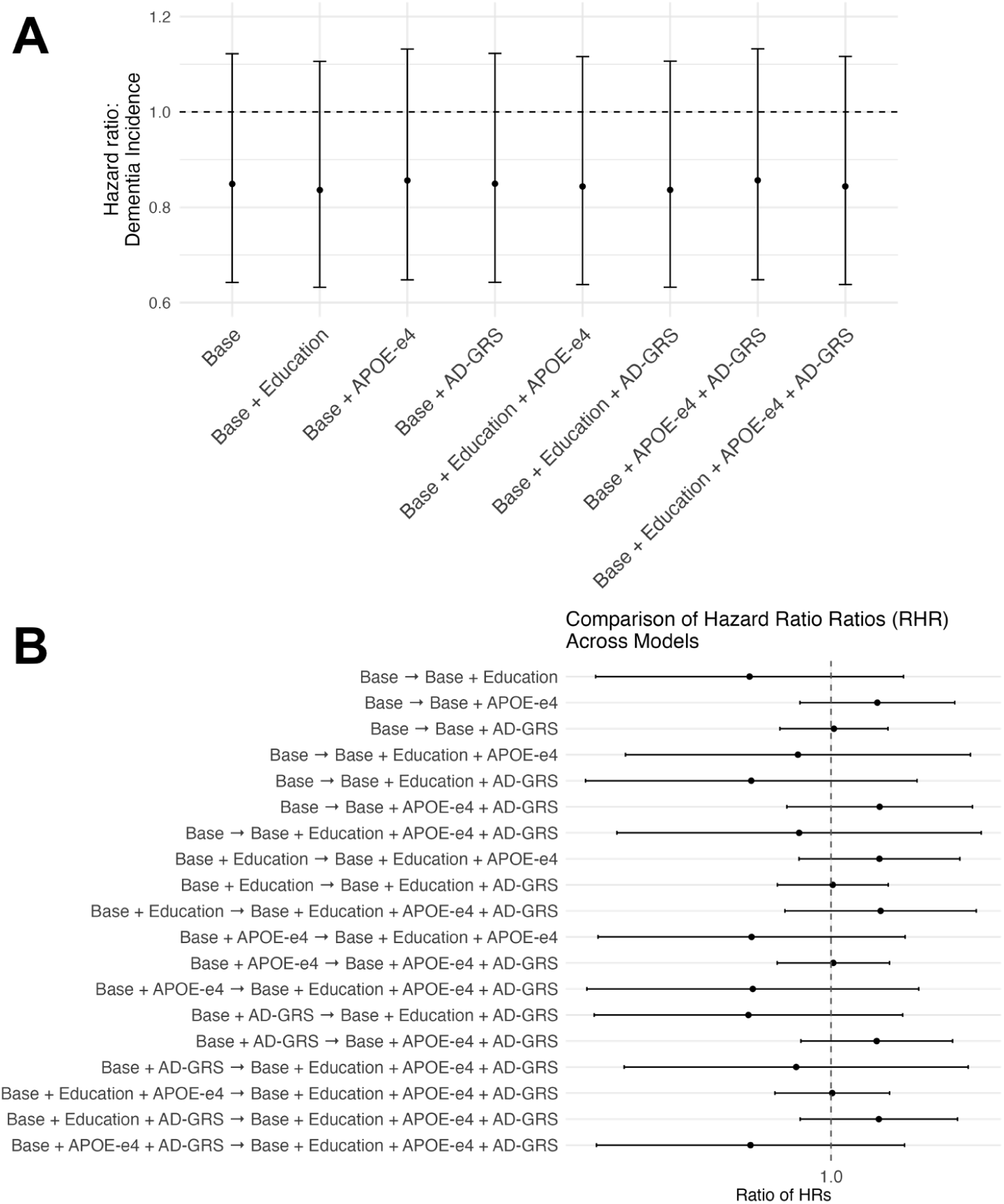
Associations between GLP-1 RA initiation versus non-SGLT2 inhibitor comparators and dementia across adjustment sets. (A) Hazard ratios for dementia incidence comparing GLP-1 RA initiatiors with comparator therapies across progressively adjusted Cox proportional hazards models, including all combinations of confounders that are typically not adjusted for: education, APOE-ε4 genotype, and AD-GRS. (B) Ratios of hazard ratios (RHRs) comparing effect estimates between models, quantifying the relative change in the GLP-1-dementia association after additional covariate adjustment with typically unadjusted-for confounders. Points indicate point estimates and lines indicate 95% confidence intervals; the vertical dashed line denotes no change (RHR = 1).

## Conclusions

Using data from the All of Us study with an active-comparator, new-user design, we found that potential confounders that are typically unavailable in EHR data cannot account for observed discrepancies between trial findings and observational associations for GLP-1 RA initiation and incident dementia. First, we found that adjustment for education and AD-related genetic risk resulted in minimal attenuation of treatment-outcome associations. Second, hazard ratios we estimated closely align with those reported in RCTs, rather than with the more optimistic estimates commonly reported in published observational studies.^3,6,11,16,20^ Because our analyses were deliberately designed to mirror existing target-trial emulations with respect to confounder adjustment and estimands, this pattern raises the possibility that cohort-related differences (i.e., effect measure modifier distribution differences) may contribute to the discrepancy between observational and RCT findings. Finally, *APOE*-ε4 carriers were substantially less likely to initiate treatment with GLP-1 RAs. This finding has important implications for research on these drugs for dementia and cardiovascular outcomes, since *APOE*-ε4 is a determinant of both.

Although education and AD genetic risk did not explain previously reported protective associations between GLP-1 RA initiation and dementia, residual confounding may still account for discrepancies between observational studies and randomized trials. Given socially patterned use and drug shortages,^38,39^ confounding in GLP-1 RA-dementia associations could be stronger than for other drugs. While higher education strongly predicted greater GLP-1 RA initiation, it did not explain these discrepancies. The association of education with dementia incidence in this cohort was modest despite being a well-established population-level risk factor.^40^ This may reflect the relatively limited follow-up, selection into the EHR-linked sample, or outcome misclassification inherent to EHR data. The attenuated education-dementia association in this data may partly explain why adjustment for education did not change treatment effect estimates. In addition, education may incompletely capture socioeconomic status (SES). Other dimensions of SES that are often unavailable or inconsistently measured in EHR-based studies—such as income, wealth, insurance coverage, and neighborhood deprivation—could contribute to residual confounding. Although we did not evaluate these factors directly, they remain important considerations for future observational research examining GLP-1 RA initiation and dementia risk.

This study has the following strengths. First, All of Us is a particularly valuable study for considering the implications of unmeasured confounding in observational research.^27^ It combines survey, genetic, and EHR data, making it possible to control for factors that are difficult to control for in most real-world data sources. While UK Biobank also combines EHR, genetic, and survey data, GLP-1 RA uptake is slower in the United Kingdom than in the US.^41^ Second, we employed an active-comparator, new-user design, aligning follow-up at treatment initiation and reducing confounding by indication and time-related biases. The use of two clinically relevant comparator groups further allows us to assess whether findings were consistent across therapeutic contrasts. Finally, our effect estimates closely aligned with those observed in randomized trials, and stability analyses yielded consistent findings, supporting the robustness of our results.

This study has the following limitations. First, we did not employ causal survival analysis or methods accounting for competing events. Although Cox proportional-hazards models have limitations in the presence of competing risks and for causal interpretation, their use here is consistent with extant approaches in the literature.^13,14,19^ Second, All of Us does not have the sample size of large EHR databases.^42^ Since dementia is a rare outcome, we had to merge users GLP1 and GIP/GLP-1 RAs together, and these drugs may not have identical effects on dementia risk. The relatively small number of incident dementia events resulted in wide confidence intervals, limiting precision for both the treatment-dementia associations and the assessment of changes after additional covariate adjustment. However, to date, there is no credible RCT evidence that effects differ by drug or that any single GLP-1 RA is effective for reducing dementia risk. Furthermore, our goal was not to assess the effect of GLP-1 RAs on dementia, but to assess the potential for unmeasured confounding by these factors. Finally, we did not rule out potential unmeasured confounding by other factors. Specifically, there is potential residual confounding related to the distinct clinical context in which SGLT2 inhibitors are prescribed, including heart failure, chronic kidney disease, and concerns about volume management.

Among adults with type 2 diabetes, additional adjustment for measured educational attainment and Alzheimer disease genetic susceptibility produced little change in estimated associations between GLP-1 RA initiation and incident dementia. These findings suggest that these measured factors were not major drivers of the estimated association in this cohort, but they do not exclude confounding by these factors in other settings or residual confounding from other sources. Selection, measurement, treatment timing, follow-up, and differences in study populations and design remain important considerations when interpreting the observational literature. The findings should not be interpreted as establishing dementia prevention as an indication for GLP-1 RA treatment.

## GenAI Disclosure

ChatGPT-4 and 5 models were used for code debugging and minor language editing to improve concision to meet the word limit. All outputs were reviewed by multiple authors, who take full responsibility for the final manuscript.

## References

1. Yao H, Zhang A, Li D, et al. Comparative effectiveness of GLP-1 receptor agonists on glycaemic control, body weight, and lipid profile for type 2 diabetes: systematic review and network meta-analysis. BMJ. 2024;384:e076410. doi:10.1136/bmj-2023-076410

2. Wilding JPH, Batterham RL, Calanna S, et al. Once-Weekly Semaglutide in Adults with Overweight or Obesity. N Engl J Med. 2021;384(11):989–1002. doi:10.1056/NEJMoa2032183

3. Kong F, Zhao Y, Zhang W, et al. Comprehensive evaluation of GLP-1 receptor agonists: an umbrella review of clinical outcomes across multiple diseases. Nat Commun. 2026;17(1):972. doi:10.1038/s41467-025-67701-9

4. Kopp KO, Glotfelty EJ, Li Y, Greig NH. Glucagon-like peptide-1 (GLP-1) receptor agonists and neuroinflammation: Implications for neurodegenerative disease treatment. Pharmacological Research. 2022;186:106550. doi:10.1016/j.phrs.2022.106550

5. Wong CK, Drucker DJ. Antiinflammatory actions of glucagon-like peptide-1–based therapies beyond metabolic benefits. J Clin Invest. 2025;135(21). doi:10.1172/JCI194751

6. Cukierman-Yaffe T, Gerstein HC, Colhoun HM, et al. Effect of dulaglutide on cognitive impairment in type 2 diabetes: an exploratory analysis of the REWIND trial. The Lancet Neurology. 2020;19(7):582–590. doi:10.1016/S1474-4422(20)30173-3

7. Nørgaard CH, Friedrich S, Hansen CT, et al. Treatment with glucagon-like peptide-1 receptor agonists and incidence of dementia: Data from pooled double-blind randomized controlled trials and nationwide disease and prescription registers. Alzheimers Dement (N Y). 2022;8(1):e12268. doi:10.1002/trc2.12268

8. Edison P, Femminella GD, Ritchie C, et al. Liraglutide in mild to moderate Alzheimer’s disease: a phase 2b clinical trial. Nat Med. 2026;32(1):353–361. doi:10.1038/s41591-025-04106-7

9. Atri A, Feldman HH, Hansen CT, et al. evoke and evoke+: design of two large-scale, double-blind, placebo-controlled, phase 3 studies evaluating the neuroprotective effects of semaglutide in early Alzheimer’s disease. Alzheimer’s & Dementia. 2022;18(S10):e062415. doi:10.1002/alz.062415

10. Cummings JL, Atri A, Feldman HH, et al. evoke and evoke+: design of two large-scale, double-blind, placebo-controlled, phase 3 studies evaluating efficacy, safety, and tolerability of semaglutide in early-stage symptomatic Alzheimer’s disease. Alzheimer’s Research & Therapy. 2025;17(1):14. doi:10.1186/s13195-024-01666-7

11. CTAD 2025 - evoke and evoke+: Two phase 3 randomised placebo-controlled trials of semaglutide in participants with early-stage Alzheimer’s disease (NCT04777396 and NCT04777409). Accessed February 20, 2026. https://sciencehub.novonordisk.com/congresses/ctad2025/johannsen1.html

12. Cummings JL, Atri A, Sano M, et al. Efficacy and safety of oral semaglutide 14 mg (flexible dose) in early-stage symptomatic Alzheimer’s disease (evoke and evoke+): two phase 3, randomised, placebo-controlled trials. The Lancet. 2026;407(10544):2167–2179. doi:10.1016/S0140-6736(26)00459-9

13. Tang B, Sjölander A, Wastesson JW, et al. Comparative effectiveness of glucagon-like peptide-1 agonists, dipeptidyl peptidase-4 inhibitors, and sulfonylureas on the risk of dementia in older individuals with type 2 diabetes in Sweden: an emulated trial study. eClinicalMedicine. 2024;73:102689. doi:10.1016/j.eclinm.2024.102689

14. Wu JY, Lin YM, Hsu WH, et al. Glucagon-Like Peptide-1 Receptor Agonists and Dementia Risk Reduction in Older Adults With Type 2 Diabetes: A Retrospective Cohort Study. J Am Med Dir Assoc. Published online October 8, 2025:105901. doi:10.1016/j.jamda.2025.105901

15. Inoue K, Saliba D, Gotanda H, et al. Glucagon-Like Peptide-1 Receptor Agonists and Incidence of Dementia Among Older Adults With Type 2 Diabetes. Ann Intern Med. 2025;178(9):1258–1267. doi:10.7326/ANNALS-24-02648

16. Sun M, Wang X, Lu Z, et al. Comparative effectiveness of SGLT2 inhibitors and GLP-1 receptor agonists in preventing Alzheimer’s disease, vascular dementia, and other dementia types among patients with type 2 diabetes. Diabetes Metab. 2025;51(2):101623. doi:10.1016/j.diabet.2025.101623

17. Tang H, Donahoo WT, DeKosky ST, et al. GLP-1RA and SGLT2i Medications for Type 2 Diabetes and Alzheimer Disease and Related Dementias. JAMA Neurol. 2025;82(5):439–449. doi:10.1001/jamaneurol.2025.0353

18. Wang W, Wang Q, Qi X, et al. Associations of semaglutide with first-time diagnosis of Alzheimer’s disease in patients with type 2 diabetes: Target trial emulation using nationwide real-world data in the US. Alzheimer’s & Dementia. 2024;20(12):8661–8672. doi:10.1002/alz.14313

19. Giorgi RD, Koychev I, Adler AI, et al. 12-month neurological and psychiatric outcomes of semaglutide use for type 2 diabetes: a propensity-score matched cohort study. eClinicalMedicine. 2024;74. doi:10.1016/j.eclinm.2024.102726

20. DiGiovanni A, Shehaj A, Millar D, Tse C, Rizk E. Utility of Pharmacological Agents for Diabetes Mellitus in the Prevention of Alzheimer’s Disease: Comparison of Metformin, Glucagon-Like Peptide-1 (GLP-1) Agonists, Insulin, and Sulfonylureas. Cureus. 2025;17(7):e87350. doi:10.7759/cureus.87350

21. Eberly LA, Yang L, Essien UR, et al. Racial, Ethnic, and Socioeconomic Inequities in Glucagon-Like Peptide-1 Receptor Agonist Use Among Patients With Diabetes in the US. JAMA Health Forum. 2021;2(12):e214182. doi:10.1001/jamahealthforum.2021.4182

22. Nguyen TT, Tchetgen EJT, Kawachi I, et al. Instrumental variable approaches to identifying the causal effect of educational attainment on dementia risk. Annals of Epidemiology. 2016;26(1):71–76.e3. doi:10.1016/j.annepidem.2015.10.006

23. Brenowitz WD, Zimmerman SC, Filshtein TJ, et al. Extension of Mendelian Randomization to Identify Earliest Manifestations of Alzheimer Disease: Association of Genetic Risk Score for Alzheimer Disease With Lower Body Mass Index by Age 50 Years. American Journal of Epidemiology. 2021;190(10):2163–2171. doi:10.1093/aje/kwab103

24. Yuan S, Wu W, Ma W, et al. Body mass index, genetic susceptibility, and Alzheimer’s disease: a longitudinal study based on 475,813 participants from the UK Biobank. J Transl Med. 2022;20(1):417. doi:10.1186/s12967-022-03621-2

25. Lumsden AL, Mulugeta A, Zhou A, Hyppönen E. Apolipoprotein E (APOE) genotype-associated disease risks: a phenome-wide, registry-based, case-control study utilising the UK Biobank. eBioMedicine. 2020;59:102954. doi:10.1016/j.ebiom.2020.102954

26. Korologou-Linden R, Bhatta L, Brumpton BM, et al. The causes and consequences of Alzheimer’s disease: phenome-wide evidence from Mendelian randomization. Nat Commun. 2022;13(1):4726. doi:10.1038/s41467-022-32183-6

27. Goldstein ND, Olivieri-Mui B, Burstyn I. Are Aggregated Electronic Health Record Datasets Good for Research? J GEN INTERN MED. 2025;40(15):3743–3749. doi:10.1007/s11606-025-09808-9

28. Ray WA. Evaluating Medication Effects Outside of Clinical Trials: New-User Designs. Am J Epidemiol. 2003;158(9):915–920. doi:10.1093/aje/kwg231

29. Lund JL, Richardson DB, Stürmer T. The Active Comparator, New User Study Design in Pharmacoepidemiology: Historical Foundations and Contemporary Application. Curr Epidemiol Rep. 2015;2(4):221–228. doi:10.1007/s40471-015-0053-5

30. Stürmer T, Wang T. Active Comparator New User Cohort Studies and Matching. JAMA Intern Med. 2026;186(1):122–123. doi:10.1001/jamainternmed.2025.5792

31. The “All of Us” Research Program | New England Journal of Medicine. Accessed February 19, 2025. https://www.nejm.org/doi/full/10.1056/NEJMsr1809937

32. Ramirez AH, Sulieman L, Schlueter DJ, et al. The All of Us Research Program: Data quality, utility, and diversity. PATTER. 2022;3(8). doi:10.1016/j.patter.2022.100570

33. Teo YH, Teo YN, Syn NL, et al. Effects of Sodium/Glucose Cotransporter 2 (SGLT2) Inhibitors on Cardiovascular and Metabolic Outcomes in Patients Without Diabetes Mellitus: A Systematic Review and Meta-Analysis of Randomized-Controlled Trials. J Am Heart Assoc. 2021;10(5):e019463. doi:10.1161/JAHA.120.019463

34. Kunkle BW, Grenier-Boley B, Sims R, et al. Genetic meta-analysis of diagnosed Alzheimer’s disease identifies new risk loci and implicates Aβ, tau, immunity and lipid processing. Nat Genet. 2019;51(3):414–430. doi:10.1038/s41588-019-0358-2

35. Choi M, Zimmerman SC, Buto PT, et al. Association of genetic risk score for Alzheimer’s disease with late-life body mass index in all of us: Evaluating reverse causation. Alzheimer’s & Dementia. 2025;21(4):e14598. doi:10.1002/alz.14598

36. Zou G. A Modified Poisson Regression Approach to Prospective Studies with Binary Data. Am J Epidemiol. 2004;159(7):702–706. doi:10.1093/aje/kwh090

37. Wang J, Choi M, Buto P, et al. Detection Bias in EHR-Based Research on Clinical Exposures and Dementia. JAMA Netw Open. 2025;8(4):e256637–e256637. doi:10.1001/jamanetworkopen.2025.6637

38. Eberly LA, Yang L, Essien UR, et al. Racial, Ethnic, and Socioeconomic Inequities in Glucagon-Like Peptide-1 Receptor Agonist Use Among Patients With Diabetes in the US. JAMA Health Forum. 2021;2(12):e214182. doi:10.1001/jamahealthforum.2021.4182

39. Walker S, Dietze P, Higgs P, et al. Socioeconomic consequences of the COVID-19 pandemic for people who use drugs. Australian Journal of Social Issues. 2023;58(4):907–925. doi:10.1002/ajs4.289

40. Glymour MM, Kawachi I, Jencks CS, Berkman LF. Does childhood schooling affect old age memory or mental status? Using state schooling laws as natural experiments. Journal of Epidemiology & Community Health. 2008;62(6):532–537. doi:10.1136/jech.2006.059469

41. Ibrahim ARN, Orayj KM. Impact of ADA Guidelines and Medication Shortage on GLP-1 Receptor Agonists Prescribing Trends in the UK: A Time-Series Analysis with Country-Specific Insights. Journal of Clinical Medicine. 2024;13(20). doi:10.3390/jcm13206256

42. Wang J, Ferguson EL, Buto P, et al. Sociodemographic, health-related, and clinical characteristics and their associations with mortality among All of Us participants compared with the United States general population. Am J Epidemiol. 2025;194(9):2477–2488. doi:10.1093/aje/kwaf118

