## Supplemental Materials for "Education and Alzheimer Disease Genetic Risk in Associations of GLP-1 Receptor Agonists With Dementia Among Adults With Type 2 Diabetes"

Dementia subtypes

Definition of censoring date

Definition of medical centers with more consistent follow-up

Definition of Lookback Period for Diagnoses

Prescription Data

#### Supplemental Tables

eTable1. Codes for outcome, exposure, and covariates.

#### Supplemental Figures

eFigure 1. Flow diagram of inclusion criteria.

eFigure 3. Association between GLP-1 RA initiation versus non-SGLT2 inhibitor comparators and dementia across nested models in individuals initiating second line therapy at age 55+.

eFigure 4. Association between GLP-1 RA initiation versus SGLT2 inhibitor and dementia across nested models in individuals initiating second line therapy at age 55+.

eFigure 5. Association between GLP-1 RA initiation versus non-SGLT2 inhibitor comparators and dementia across nested models in the EHR subsample with better followup.

eFigure 6. Association between GLP-1 RA versus SGLT2 inhibitor use and dementia in the EHR subsample with better followup.

eFigure 7. Association between GLP-1 RA initiation versus non-SGLT2 inhibitor comparators and dementia across nested models adjusting for obesity.

eFigure 8. Association between GLP-1 RA initiation versus SGLT2 inhibitor and dementia across nested models in individuals initiating second line therapy and adjusting for obesity.

eFigure 9. Association between GLP-1 RA initiation versus non-SGLT2 inhibitor comparators and dementia across nested models in individuals with mild cognitive impairment.

eFigure 10. Association between GLP-1 RA initiation versus SGLT2 inhibitor and dementia across nested models in individuals initiating second line therapy and with mild cognitive impairment.

### Supplemental Methods

#### Dementia subtypes

We combined dementia subtypes because clinical dementia diagnoses often correspond imperfectly with underlying neuropathology. In older adults, mixed pathology is common, with Alzheimer's disease, vascular, Lewy body/α-synuclein, and other pathologies frequently co-occurring. Distinguishing subtypes based on clinical diagnosis alone may therefore introduce misclassification and reduce interpretability, whereas a combined dementia outcome better reflects the etiologic heterogeneity of dementia in real-world settings.

#### Associations of education and AD genetic risk with dementia incidence

We estimated the associations between education, AD genetic risk and dementia incidence using Cox models. Participants were followed from the baseline of the active comparator design until the earliest date of dementia diagnosis, death, or censoring.

#### Definition of censoring date

We defined censoring due to loss to follow-up as no recorded EHR encounter for 448 days, corresponding to the 99th percentile of the distribution of days between encounters. Individuals meeting this criterion were assumed to no longer be receiving care at a clinic with linked EHR data and were censored 448 days after their last recorded encounter.

#### Definition of medical centers with more consistent follow-up

We first summarized follow up characteristics at each EHR site, including the number of participants, visit counts, duration of follow up, and gaps between visits. Sites were classified as reasonable if at least 50% of participants had $\geq 3$ visits, $\geq 1$ year of follow up, and no visit gaps exceeding 2 years. We then identified all participants with at least one recorded visit at a site meeting these criteria. A participant level indicator was created to flag individuals with EHR follow up at a reasonable site for use in sensitivity analyses.

#### Definition of Lookback Period for Diagnoses

Diagnosis indicators were defined using all available diagnosis information prior to the index date. Participants were classified as having a prior diagnosis if the relevant diagnosis appeared at any time in their available record before the index date. Thus, the lookback period varied across participants according to the length of observable pre-index history.

#### Prescription Data

Medication exposure was identified from All of Us EHR prescription data using RxNorm concept identifiers. Participants were classified according to initiation of a GLP-1 receptor agonist (GLP-1 RA), sodium-glucose cotransporter-2 inhibitor (SGLT2i), or other second-line antidiabetic therapy based on the first recorded prescription after type 2 diabetes diagnosis. The treatment strategies evaluated here should be interpreted as point-treatment initiation strategies rather than sustained-use strategies; we did not consider the available data sufficient to identify the effects of sustained treatment use over time, particularly given that treatment is often interrupted or discontinued after initiation because of intercurrent clinical events.

### Supplemental Tables

eTable1. Codes for outcome, exposure, and covariates.

| Construct | ICD-9 and ICD-10 Codes | OMOP Concept/RxNorm IDs |
| --- | --- | --- |
| *Inclusion Criteria* |  |  |
| Available EHR data |  |  |
| Type 2 diabetes | **ICD-10 Codes:** E11.0, E11.1, E11.2, E11.3, E11.4, E11.5, E11.6, E11.7, E11.8, E11.9<br>**ICD-9 Codes:** 250.00, 250.02, 250.10, 250.12, 250.20, 250.22, 250.30, 250.32, 250.40, 250.42, 250.50, 250.52, 250.60, 250.62, 250.70, 250.72, 250.80, 250.82, 250.90, 250.92 | Type 2 diabetes mellitus with hyperosmolarity, Type 2 diabetes mellitus with ketoacidosis, Type 2 diabetes mellitus with kidney complications, Type 2 diabetes mellitus with ophthalmic complications, Type 2 diabetes mellitus with neurological complications, Type 2 diabetes mellitus with circulatory complications, Type 2 diabetes mellitus with other specified complications, Type 2 diabetes mellitus with multiple complications, Type 2 diabetes mellitus with unspecified complications, Type 2 diabetes mellitus without complications, Diabetes mellitus without mention of complication, type II or unspecified type, not stated as uncontrolled, Diabetes mellitus without mention of complication, type II or unspecified type, uncontrolled, Diabetes with ketoacidosis, type II or unspecified type, not stated as uncontrolled, Diabetes with ketoacidosis, type II or unspecified type, uncontrolled, Diabetes with hyperosmolarity, type II or unspecified |
|  |  | type, not stated as uncontrolled, Diabetes with hyperosmolarity, type II or unspecified type, uncontrolled, Diabetes with other coma, type II or unspecified type, not stated as uncontrolled, Diabetes with other coma, type II or unspecified type, uncontrolled, Diabetes with renal manifestations, type II or unspecified type, not stated as uncontrolled, Diabetes with renal manifestations, type II or unspecified type, uncontrolled, Diabetes with ophthalmic manifestations, type II or unspecified type, not stated as uncontrolled, Diabetes with ophthalmic manifestations, type II or unspecified type, uncontrolled, Diabetes with neurological manifestations, type II or unspecified type, not stated as uncontrolled, Diabetes with neurological manifestations, type II or unspecified type, uncontrolled, Diabetes with peripheral circulatory disorders, type II or unspecified type, not stated as uncontrolled, Diabetes with peripheral circulatory disorders, type II or unspecified type, uncontrolled, Diabetes with other specified manifestations, type II or unspecified type, not stated as uncontrolled, Diabetes with other specified manifestations, type II or unspecified type, uncontrolled, Diabetes with unspecified complication, type II or unspecified |
|  |  | type, not stated as uncontrolled,<br>Diabetes with unspecified<br>complication, type II or unspecified<br>type, uncontrolled |
| Exclusion Criteria for Active Comparator Designs<br>*Before Drug Initiation* |  |  |
| Pancreatitis | **ICD-10 Codes:** K85.0, K85.1,<br>K85.2, K85.3, K85.8, K85.9, K86.0,<br>K86.1<br>**ICD-9 Codes:** 577.0, 577.1 | Pancreatitis, Acute pancreatitis,<br>Chronic pancreatitis, Alcohol-induced<br>pancreatitis, Drug-induced acute<br>pancreatitis, Gallstone pancreatitis |
| Thyroid cancer | **ICD-10 Codes:** C73, Z85.850,<br>E31.2<br>**ICD-9 Codes:** 193, V10.87, 258.02 | Malignant tumor of thyroid gland,<br>Multiple endocrine neoplasia type II |
| Type 1 diabetes | **ICD-10 Codes:** E10.9, E10.10,<br>E10.11, E10.21, E10.29, E10.31,<br>E10.39, E10.41, E10.49, E10.51,<br>E10.52, E10.59, E10.610, E10.618,<br>E10.620, E10.621, E10.622,<br>E10.628, E10.65, E10.69, E10.8<br>**ICD-9 Codes:** 250.01, 250.03 | Type 1 diabetes mellitus, Insulin<br>dependent diabetes mellitus type 1A,<br>Insulin dependent diabetes mellitus<br>type 1B, Type 1 diabetes mellitus<br>without complication, Type 1 diabetes<br>mellitus with ulcer, Type 1 diabetes<br>mellitus with arthropathy |
| Gastroparesis | **ICD-10 Code:** K31.84<br>**ICD-9 Code:** 536.3 | Gastroparesis syndrome |
| AD medication use |  | Donepezil (RxNorm: 135447)<br>Rivastigmine (RxNorm: 183379)<br>Galantamine (RxNorm: 4637)<br>Memantine (RxNorm: 6719) |
| Traumatic Brain Injury | **ICD-10 Codes:** S06.0X0A,<br>S06.1X0A, S06.2X0A, S06.4X0A,<br>S06.5X0A, S06.6X0A, S06.9X0A<br>**ICD-9 Codes:** 850.0, 851.0, 851.1,<br>851.8, 851.9, 852.0, 852.1, 852.2, | Concussion, Traumatic cerebral<br>edema, Epidural hemorrhage,<br>Traumatic subdural hemorrhage,<br>Traumatic subarachnoid hemorrhage,<br>Traumatic brain injury |
|  | 852.3, 853.00, 854.00, V15.52 |  |
|  | *Potential Confounders (Covariates)* |  |
| Age (/birth year), sex, race/ethnicity, education, APOE-ε4, AD-GRS |  |  |
| Diabetes Complications | **ICD-10 Codes:** E10.319, E10.359, E11.319, E11.359, E10.40, E10.42, E11.40, E11.42, E10.51, E11.51, E10.610, E11.610, E10.618, E11.618, E10.69, E11.69, E11.65<br>**ICD-9 Codes:** 362.01, 362.02, 250.60, 250.61, 250.70, 250.71, 250.80, 250.81, 250.90, 250.91 | Retinopathy due to diabetes mellitus, Proliferative retinopathy due to diabetes mellitus, Neuropathy due to diabetes mellitus, Polyneuropathy due to diabetes mellitus, Peripheral vascular disease, Peripheral angiopathy due to diabetes mellitus, Hyperglycemia, Hyperglycemia due to diabetes mellitus |
| Hypertension | **ICD-10 Codes:** I10, I11.0, I11.9, I12.0, I12.9, I13.0, I13.10, I13.11, I13.2, I15.0, I15.1, I15.2, I15.8, I15.9, I16.0, I16.1, I16.9<br>**ICD-9 Codes:** 401.0, 401.1, 401.9, 402.00, 402.01, 402.10, 402.11, 402.90, 402.91, 403.00, 403.01, 403.10, 403.11, 403.90, 403.91, 404.00, 404.01, 404.02, 404.03, 404.10, 404.11, 404.12, 404.13, 404.90, 404.91, 404.92, 404.93, 405.01, 405.09, 405.11, 405.19, 405.91, 405.99 | Essential hypertension, Hypertensive heart disease, Hypertensive heart and renal disease with (congestive) heart failure, Hypertensive heart disease without congestive heart failure, Hypertensive heart and chronic kidney disease, Hypertensive chronic kidney disease stage 1 through stage 4, Hypertensive chronic kidney disease stage 5, Secondary hypertension, Renovascular hypertension, Hypertension secondary to endocrine disorder, Other secondary hypertension, Secondary hypertension, unspecified, Hypertensive urgency, Hypertensive emergency, Hypertensive crisis, unspecified, Resistant hypertension |
| Dyslipidemia and Lipoprotein Metabolism Disorders | **ICD-10 Codes:** E78.0, E78.1, E78.2, E78.4, E78.5, E78.9<br>**ICD-9 Codes:** 272.0, 272.1, 272.2, 272.4, 272.5, 272.9 | Hypercholesterolemia, Pure hyperglyceridemia, Lipoprotein deficiency, Hyperlipidemia, Mixed hyperlipidemia, Other hyperlipidemia, Hyperlipidemia, unspecified, Disorder of lipoprotein metabolism, Disorders of lipoprotein metabolism and other lipidemias |
| Cardiovascular Diseases and Heart Conditions | **ICD-10 Codes:** I20.0, I20.1, I20.8, I20.9, I21.0, I21.1, I21.2, I21.3, I21.9, I22.0, I22.1, I22.2, I22.8, I22.9, I23.0, I23.1, I23.2, I23.3, I23.4, I23.5, I23.6, I23.7, I23.8, I24.0, I24.8, I24.9, I25.0, I25.1, I25.2, I25.3, I25.4, I25.5, I25.6, I25.7, I25.8, I25.9, I30.0, I30.1, I30.8, I30.9, I31.0, I31.1, I31.2, I31.3, I31.4, I31.8, I31.9, I32.0, I33.0, I33.9, I34.0, I34.1, I34.2, I34.8, I34.9, I35.0, I35.1, I35.2, I35.8, I35.9, I36.0, I36.1, I36.8, I36.9, I37.0, I37.1, I37.8, I37.9, I38.0, I38.8, I38.9, I39.0, I39.8, I40.0, I40.1, I40.8, I40.9, I41.0, I41.8<br>**ICD-9 Codes:** 413.0, 413.1, 413.9, 410.00, 410.01, 410.10, 410.11, 410.20, 410.21, 410.30, 410.31, 410.40, 410.41, 410.50, 410.51, 410.60, 410.61, 410.70, 410.71, 410.80, 410.81, 410.90, 410.91, 411.0, 411.1, 411.81, 411.89, 412, 414.0, 414.01, 414.02, 414.03, 414.04, 414.05, 414.06, 414.07, 414.8, 414.9, 420.0, 420.9, 423.0, 423.1, | Angina pectoris, Acute myocardial infarction, Subsequent myocardial infarction, Complications following myocardial infarction, Other acute ischemic heart diseases, Chronic ischemic heart disease, Acute pericarditis, Disorder of pericardium, Pericarditis, Acute and subacute endocarditis, Nonrheumatic mitral valve disorders, Nonrheumatic aortic valve disorders, Nonrheumatic tricuspid valve disorders, Nonrheumatic pulmonary valve disorders, Endocarditis, valve unspecified, Endocarditis and heart valve disorders in diseases classified elsewhere, Acute myocarditis, Myocarditis in diseases classified elsewhere |
|  | 423.2, 423.3, 423.8, 423.9, 424.0, 424.1, 424.2, 424.3, 424.90, 424.91, 424.99, 425.0, 425.1, 425.2, 425.3, 425.4, 425.5, 425.7, 425.8, 425.9, 429.0, 429.1, 429.2, 429.3 |  |
| Cerebrovascular Disease | **ICD-10 Codes:** I60.0, I60.1, I60.2, I60.3, I60.4, I60.5, I60.6, I60.7, I60.8, I60.9, I61.0, I61.1, I61.2, I61.3, I61.4, I61.5, I61.6, I61.8, I61.9, I62.0, I62.1, I62.9, I63.0, I63.1, I63.2, I63.3, I63.4, I63.5, I63.6, I63.8, I63.9, I65.0, I65.1, I65.2, I65.3, I65.8, I65.9, I66.0, I66.1, I66.2, I66.3, I66.8, I66.9, I67.0, I67.1, I67.2, I67.3, I67.4, I67.5, I67.6, I67.7, I67.8, I67.9, I68.0, I68.1, I68.2, I68.8, I69.0, I69.1, I69.2, I69.3, I69.4, I69.8<br>**ICD-9 Codes:** 430, 431, 432.0, 432.1, 432.9, 433.00, 433.01, 433.10, 433.11, 433.20, 433.21, 433.30, 433.31, 433.80, 433.81, 433.90, 433.91, 434.00, 434.01, 434.10, 434.11, 434.90, 434.91, 435.0, 435.1, 435.2, 435.3, 435.8, 435.9, 436, 437.0, 437.1, 437.2, 437.3, 437.4, 437.5, 437.6, 437.7, 437.8, 437.9, 438.0, 438.1, 438.2, 438.3, 438.4, 438.5, 438.6, 438.7, 438.8, 438.9 | Nontraumatic subarachnoid hemorrhage, Nontraumatic intracerebral hemorrhage, Other nontraumatic intracranial hemorrhage, Cerebral infarction, Occlusion and stenosis of cerebral arteries without infarction, Other cerebrovascular diseases, Cerebrovascular disorders in diseases classified elsewhere, Sequelae of cerebrovascular disease |
| Chronic Kidney | **ICD-10 Codes:** N18.1, N18.2, | Chronic kidney disease stage 1, |
| Disease and End-Stage Renal Disease | N18.30, N18.31, N18.32, N18.4, N18.5, N18.6, N18.9<br>**ICD-9 Codes:** 585.1, 585.2, 585.3, 585.4, 585.5, 585.6, 585.9, 586 | Chronic kidney disease stage 2, Chronic kidney disease stage 3, Chronic kidney disease stage 3a, Chronic kidney disease stage 3b, Chronic kidney disease stage 4, Chronic kidney disease stage 5, End stage renal disease, Chronic kidney disease, unspecified |
| Alcohol Use Disorders | **ICD-10 Codes:** F10.10, F10.11, F10.120, F10.121, F10.129, F10.130, F10.131, F10.139, F10.14, F10.150, F10.151, F10.159, F10.180, F10.181, F10.182, F10.188, F10.19, F10.20, F10.21, F10.220, F10.221, F10.229, F10.230, F10.231, F10.239, F10.24, F10.250, F10.251, F10.259, F10.280, F10.281, F10.282, F10.288, F10.29<br>**ICD-9 Codes:** 303.00, 303.01, 303.02, 303.03, 303.90, 303.91, 305.00, 305.01, 305.02, 305.03, 291.0, 291.1, 291.2, 291.3, 291.4, 291.5, 291.81, 291.82, 291.89, 291.9 | Alcohol abuse, Alcohol dependence, Alcohol use disorder, Alcohol withdrawal, Alcohol intoxication, Alcohol-induced mood disorder, Alcohol-induced psychotic disorder, Alcohol-related disorder in remission |
| History of Tobacco Use | **ICD-10 Codes:** F17.200, F17.201, F17.203, F17.208, F17.209, F17.210, F17.211, F17.213, F17.218, F17.219, F17.220, F17.221, F17.223, F17.228, F17.229, F17.290, F17.291, F17.293, F17.298, F17.299, Z87.891<br>**ICD-9 Codes:** 305.1, V15.82 | Nicotine dependence, Tobacco use disorder, Cigarette dependence, Smokeless tobacco dependence, Nicotine withdrawal, History of tobacco use |
| Depression | **ICD-10 Codes:** F32.0, F32.1, F32.2, F32.3, F32.4, F32.5, F32.8, F32.9<br>**ICD-9 Codes:** 296.20, 296.21, 296.22, 296.23, 296.24, 296.25, 296.26, 296.30, 296.31, 296.32, 296.33, 296.34, 296.35, 296.36 | Depressive disorder, mild,<br>Depressive disorder, moderate,<br>Depressive disorder, severe without psychotic features,<br>Psychotic major depression,<br>Major depression in partial remission,<br>Major depression in full remission,<br>Other specified depressive episodes,<br>Major depressive disorder, single episode |
| Sleep Disorders | **ICD-10 Codes:** G47.00, G47.01, G47.09, G47.10, G47.11, G47.19, G47.20, G47.21, G47.22, G47.23, G47.24, G47.25, G47.26, G47.27, G47.33, G47.34, G47.35, G47.36, G47.37, G47.4, G47.50, G47.51, G47.52, G47.53, G47.59, G47.8, G47.9<br>**ICD-9 Codes:** 307.41, 307.42, 307.44, 307.45, 327.00, 327.01, 327.02, 327.09, 327.10, 327.11, 327.12, 327.13, 327.14, 327.15, 327.19, 327.20, 327.21, 327.22, 327.23, 327.24, 327.25, 327.26, 327.27, 327.29, 327.30, 327.31, 327.32, 327.33, 327.34, 327.35, 327.36, 327.37, 327.39, 780.50, 780.51, 780.52, 780.53, 780.54, 780.55, 780.56, 780.57, 780.58, 780.59 | Insomnia, Hypersomnia, Disorder of sleep-wake cycle, Sleep apnea, Narcolepsy, Parasomnia,<br>Sleep-related movement disorder,<br>Other sleep disorder, Sleep disorder, unspecified |
| Liver Disease | **ICD-10 Codes:** K76.0, K75.81, K70.0, K70.1, K70.2, K70.3, K70.4, K74.0, K74.1, K74.2, K74.3, K74.4, K74.5, K74.6, K76.9<br>**ICD-9 Codes:** 571.0, 571.1, 571.2, | Nonalcoholic fatty liver disease,<br>Nonalcoholic steatohepatitis, Alcoholic liver disease, Alcoholic fatty liver,<br>Alcoholic hepatitis, Alcoholic cirrhosis of liver, Alcoholic hepatic failure, |
|  | 571.3, 571.40, 571.41, 571.42, 571.49, 571.5, 571.6, 571.8, 571.9 | Cirrhosis of liver, Liver disease |
| Sensory Impairment | **ICD-10 Codes:** H90.0, H90.11, H90.12, H90.2, H90.3, H90.41, H90.42, H90.5, H90.6, H90.7, H91.0, H91.01, H91.02, H91.03, H91.1, H91.2, H91.20, H91.21, H91.22, H91.23, H91.8X1, H91.8X2, H91.8X3, H91.8X9, H91.90, H91.91, H91.92, H91.93<br>**ICD-9 Codes:** 389.00, 389.01, 389.02, 389.03, 389.04, 389.05, 389.06, 389.08, 389.10, 389.11, 389.12, 389.14, 389.15, 389.16, 389.17, 389.18, 389.20, 389.21, 389.22, 389.8, 389.9 | Conductive hearing loss, Sensorineural hearing loss, Mixed conductive and sensorineural hearing loss, Unspecified hearing loss, Deafness, Nerve deafness, Sudden hearing loss, Noise-induced hearing loss, High frequency hearing loss, Low frequency hearing loss, Bilateral hearing loss, Unilateral hearing loss |
| *Medication Use at Baseline* |  |  |
| Insulin Use at Baseline |  | Insulin lispro (RxNorm: 86009), Insulin aspart (RxNorm: 51428), Insulin glulisine (RxNorm: 400008), Regular human insulin (RxNorm: 253182), NPH insulin (HCPCS: S5552), Insulin detemir (RxNorm: 139825), Insulin glargine (RxNorm: 274783), Insulin degludec (RxNorm: 1670007), Insulin aspart protamine/insulin aspart (RxNorm: 352385), Insulin lispro protamine/insulin lispro (RxNorm: 314684) |
| Metformin Use at Baseline |  | Metformin (RxNorm: 6809) |
| *Outcomes* |  |  |
| GLP-1 Initiation |  | Exenatide (RxNorm: 60548),<br>Liraglutide (RxNorm: 475968),<br>Dulaglutide (RxNorm: 1551291),<br>Lixisenatide (RxNorm: 1440051),<br>Semaglutide (RxNorm: 1792776)<br>Tirzepatide (RxNorm: 2479358) |
| SGLT2i Initiation |  | Canagliflozin (RxNorm: 1424323),<br>Dapagliflozin (RxNorm: 1424324),<br>Empagliflozin (RxNorm: 1545658),<br>Ertugliflozin (RxNorm: 1912215) |
| Other Second-Line<br>Therapy for Type 2<br>Diabetes:<br>dpp-4 inhibitors,<br>sulfonylureas,<br>meglitinides,<br>thiazolidinediones,<br>alpha-glucosidase<br>inhibitors, amylin<br>analog, bile acid<br>sequestrant,<br>dopamine agonist |  | Sitagliptin (RxNorm: 593411),<br>Saxagliptin (RxNorm: 857974),<br>Linagliptin (RxNorm: 1100699),<br>Alogliptin (RxNorm: 1368001),<br>Vildagliptin (RxNorm:), Gemigliptin<br>(RxNorm:), Trelagliptin (RxNorm:),<br>Glipizide (RxNorm: 4821), Glyburide<br>(RxNorm: 4815), Glimepiride<br>(RxNorm: 25789), Repaglinide<br>(RxNorm: 73044), Nateglinide<br>(RxNorm: 274332), Pioglitazone<br>(RxNorm: 33738), Rosiglitazone<br>(RxNorm: 84108), Acarbose (RxNorm:<br>16681), Miglitol (RxNorm: 30009),<br>Pramlintide (RxNorm: 139953),<br>Colesevelam (RxNorm: 141626),<br>Bromocriptine (RxNorm: 1760) |
| All-Cause Dementia | **ICD-10 Codes:** G30.0, G30.1,<br>G30.8, G30.9, G31.83, F01.50,<br>F01.51, F03.90, F03.91, F02.80,<br>F02.81<br>**ICD-9 Codes:** 331.0, 331.82,<br>290.40, 290.41, 290.42, 290.43,<br>294.20, 294.21, 294.10, 294.11 | Dementia, Diffuse Lewy body disease,<br>Alzheimer's disease, Unspecified<br>dementia, Vascular dementia,<br>Dementia in other diseases classified<br>elsewhere |

### Supplemental Figures

eFigure 1. Flow diagram of inclusion criteria.

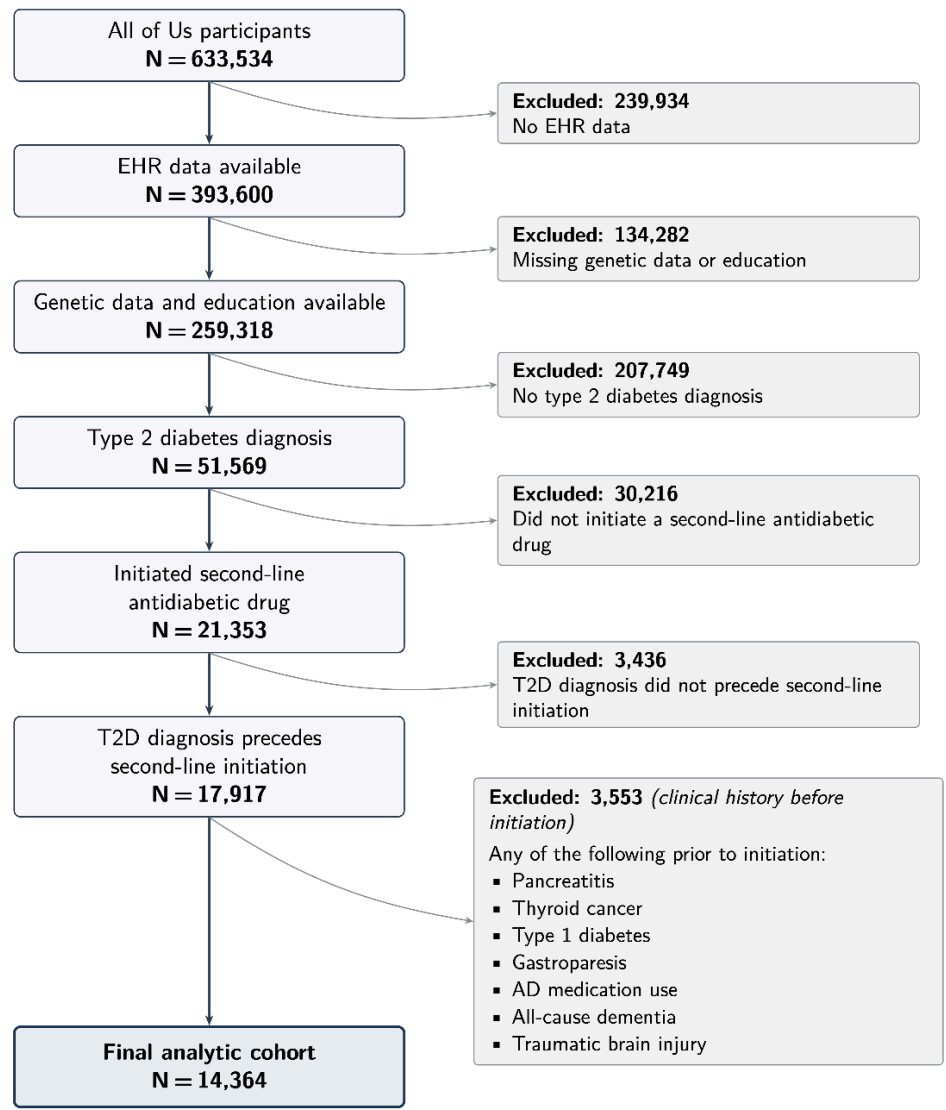

eFigure 2. Association between GLP-1 RA versus SGLT2 inhibitor use and dementia.

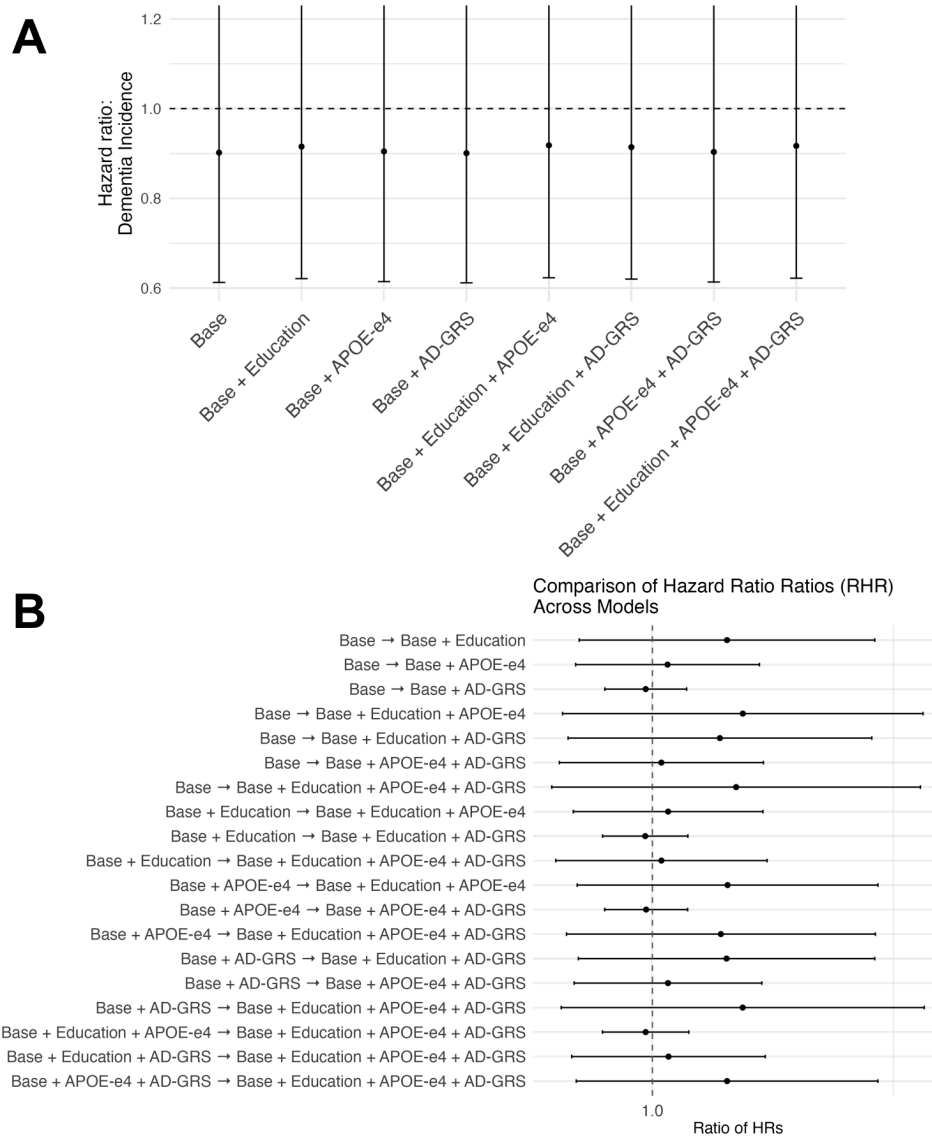

(A) Hazard ratios for dementia incidence comparing GLP-1 users with SGL2i therapies across progressively adjusted Cox proportional hazards models, including all combinations of confounders that are typically not adjusted for: education, APOE-ε4 genotype, and AD-GRS. (B) Ratios of hazard ratios (RHRs) comparing effect estimates between models, quantifying the relative change in the GLP-1-dementia association after additional covariate adjustment with typically unadjusted for confounders. Points indicate point estimates and lines indicate 95% confidence intervals; the vertical dashed line denotes no change (RHR = 1).

eFigure 3. Association between GLP-1 RA initiation versus non-SGLT2 inhibitor comparators and dementia across nested models in individuals initiating second line therapy at age 55+.

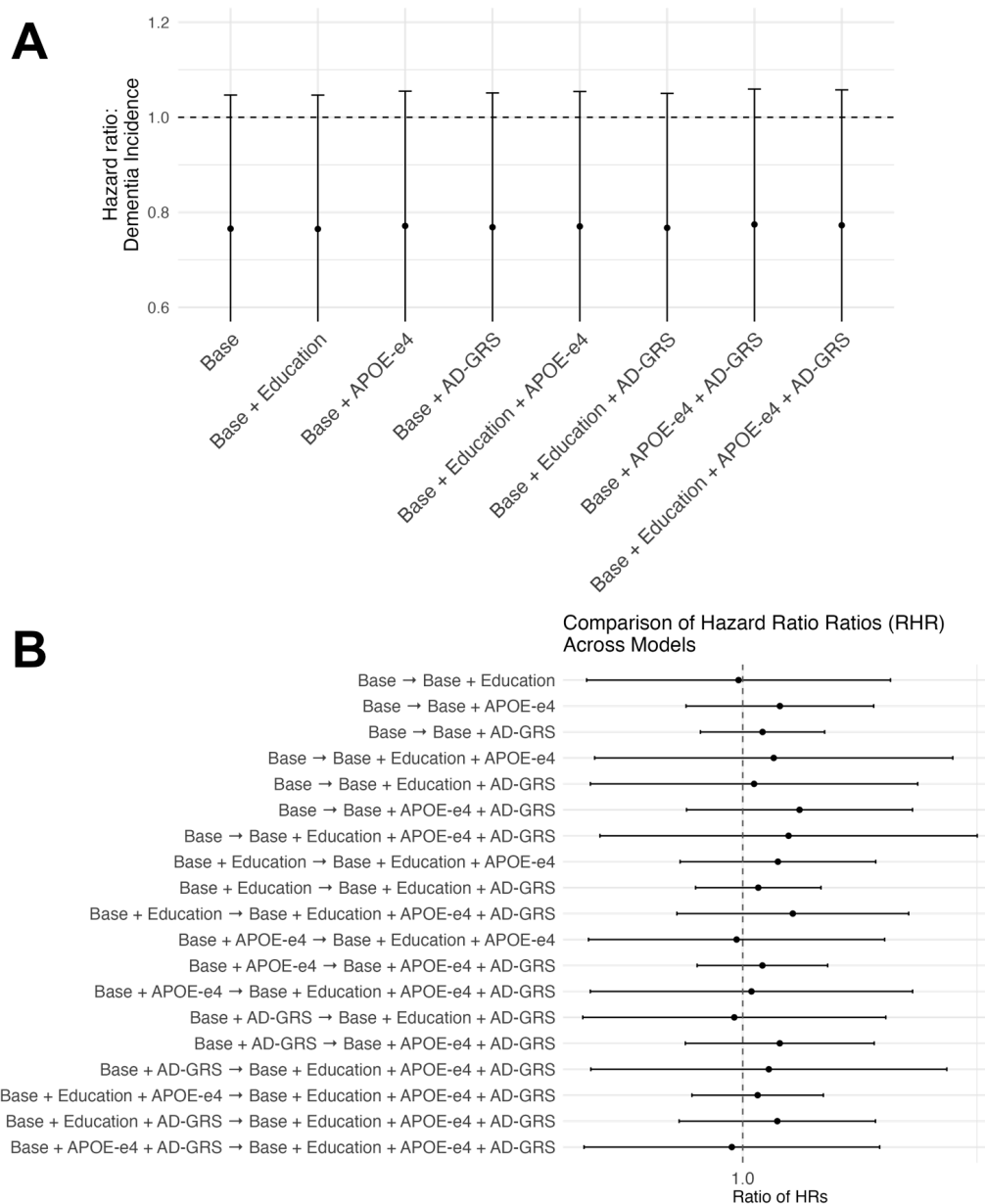

(A) Hazard ratios for dementia incidence comparing GLP-1 users with comparator therapies across progressively adjusted Cox proportional hazards models, including all combinations of confounders that are typically not adjusted for: education, APOE-ε4 genotype, and AD-GRS. (B) Ratios of hazard ratios (RHRs) comparing effect estimates between models, quantifying the relative change in the GLP-1-dementia association after additional covariate adjustment with typically unadjusted-for confounders. Points indicate point estimates and lines indicate 95% confidence intervals; the vertical dashed line denotes no change (RHR = 1).

eFigure 4. Association between GLP-1 RA initiation versus SGLT2 inhibitor and dementia across nested models in individuals initiating second line therapy at age 55+.

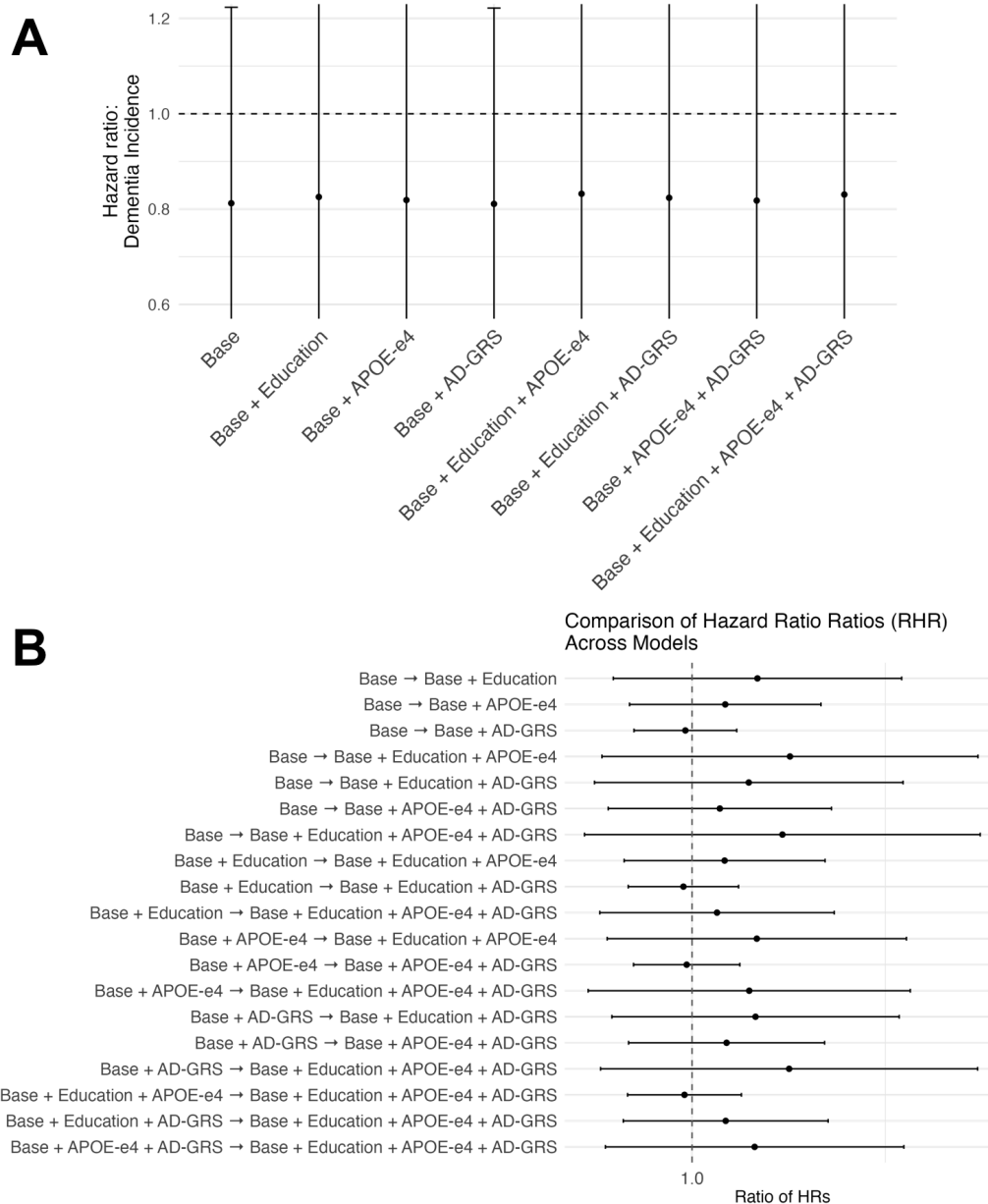

(A) Hazard ratios for dementia incidence comparing GLP-1 users with SGLT2 inhibitor use across progressively adjusted Cox proportional hazards models, including all combinations of confounders that are typically not adjusted for: education, APOE-ε4 genotype, and AD-GRS. (B) Ratios of hazard ratios (RHRs) comparing effect estimates between models, quantifying the relative change in the GLP-1-dementia association after additional covariate adjustment with typically unadjusted-for confounders. Points indicate point estimates and lines indicate 95% confidence intervals; the vertical dashed line denotes no change (RHR = 1).

eFigure 5. Association between GLP-1 RA initiation versus non-SGLT2 inhibitor comparators and dementia across nested models in the EHR subsample with better followup.

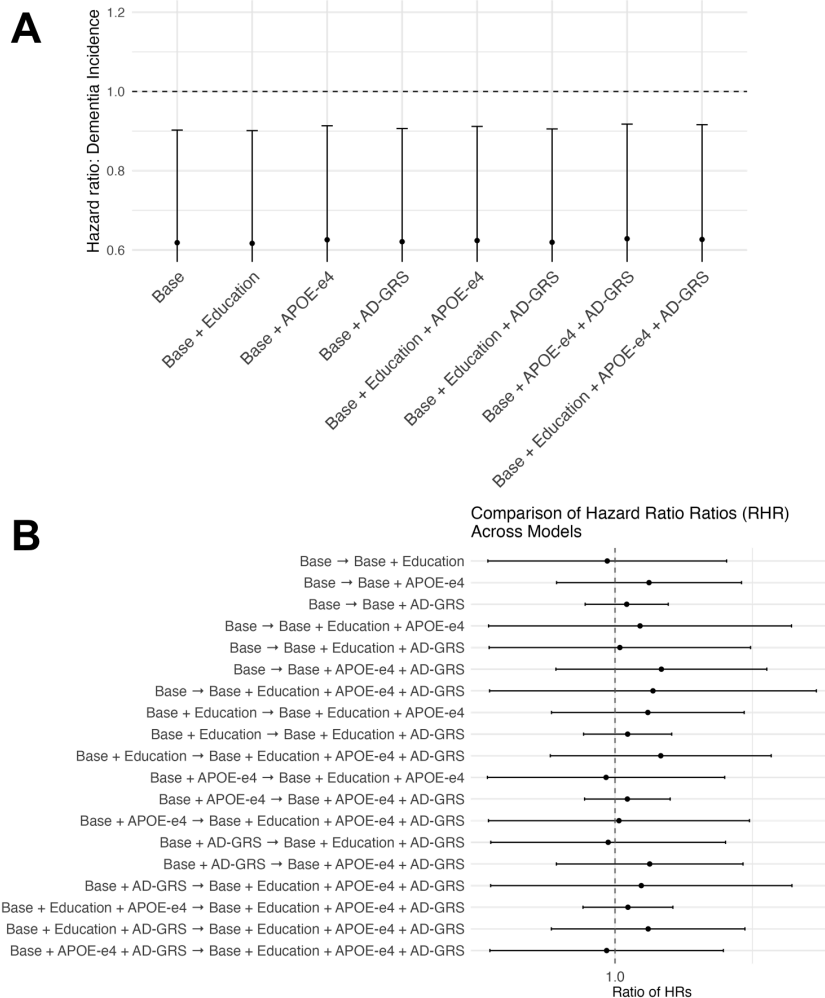

(A) Hazard ratios for dementia incidence comparing GLP-1 users with comparator therapies across progressively adjusted Cox proportional hazards models, including all combinations of confounders that are typically not adjusted for: education, APOE-ε4 genotype, and AD-GRS. (B) Ratios of hazard ratios (RHRs) comparing effect estimates between models, quantifying the relative change in the GLP-1-dementia association after additional covariate adjustment with typically unadjusted-for confounders. Points indicate point estimates and lines indicate 95% confidence intervals; the vertical dashed line denotes no change (RHR = 1).

eFigure 6. Association between GLP-1 RA versus SGLT2 inhibitor use and dementia in the EHR subsample with better followup.

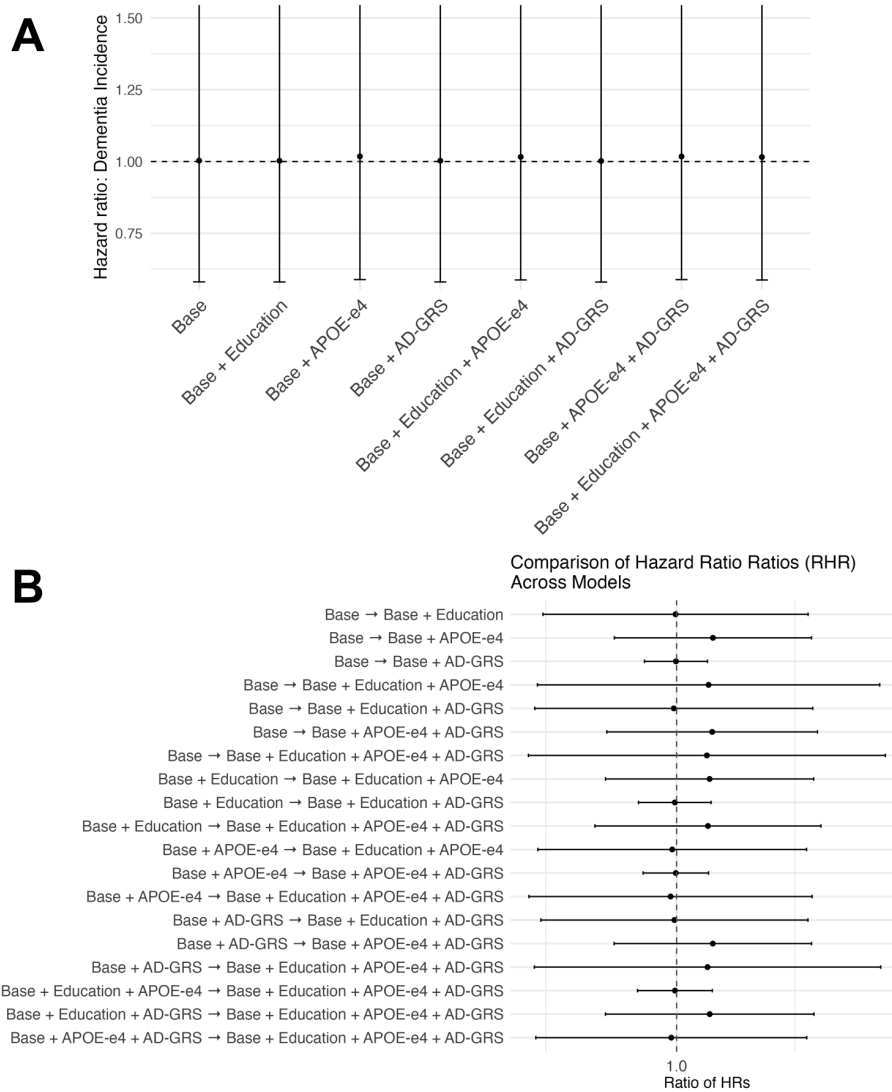

(A) Hazard ratios for dementia incidence comparing GLP-1 users with SGLT2 inhibitor use across progressively adjusted Cox proportional hazards models, including all combinations of confounders that are typically not adjusted for: education, APOE-ε4 genotype, and AD-GRS. (B) Ratios of hazard ratios (RHRs) comparing effect estimates between models, quantifying the relative change in the GLP-1-dementia association after additional covariate adjustment with typically unadjusted-for confounders. Points indicate point estimates and lines indicate 95% confidence intervals; the vertical dashed line denotes no change (RHR = 1).

eFigure 7. Association between GLP-1 RA initiation versus non-SGLT2 inhibitor comparators and dementia across nested models adjusting for obesity.

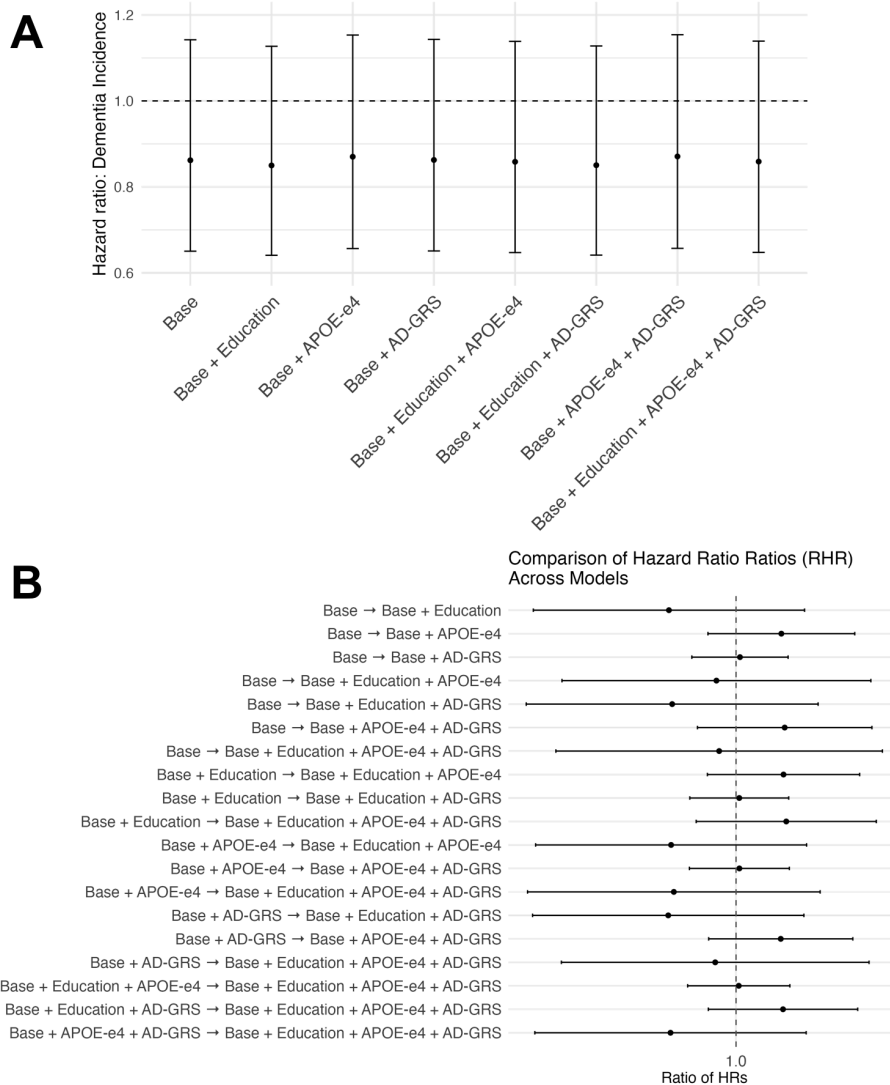

(A) Hazard ratios for dementia incidence comparing GLP-1 users with comparator therapies across progressively adjusted Cox proportional hazards models, including all combinations of confounders that are typically not adjusted for: education, APOE-ε4 genotype, and AD-GRS. (B) Ratios of hazard ratios (RHRs) comparing effect estimates between models, quantifying the relative change in the GLP-1-dementia association after additional covariate adjustment with typically unadjusted-for confounders. Points indicate point estimates and lines indicate 95% confidence intervals; the vertical dashed line denotes no change (RHR = 1).

eFigure 8. Association between GLP-1 RA initiation versus SGLT2 inhibitor and dementia across nested models in individuals initiating second line therapy and adjusting for obesity.

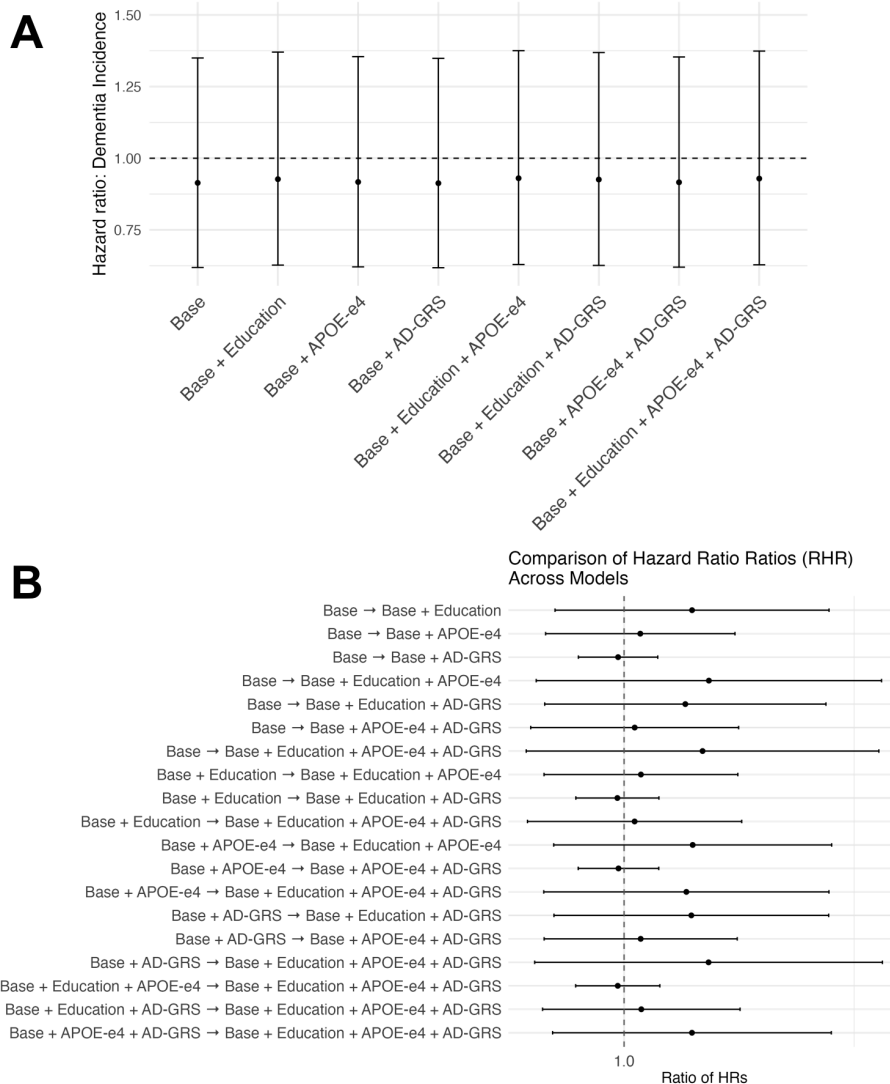

(A) Hazard ratios for dementia incidence comparing GLP-1 users with SGLT2 inhibitor use across progressively adjusted Cox proportional hazards models, including all combinations of confounders that are typically not adjusted for: education, APOE-ε4 genotype, and AD-GRS. (B) Ratios of hazard ratios (RHRs) comparing effect estimates between models, quantifying the relative change in the GLP-1-dementia association after additional covariate adjustment with typically unadjusted-for confounders. Points indicate point estimates and lines indicate 95% confidence intervals; the vertical dashed line denotes no change (RHR = 1).

eFigure 9. Association between GLP-1 RA initiation versus non-SGLT2 inhibitor comparators and dementia across nested models in individuals with mild cognitive impairment.

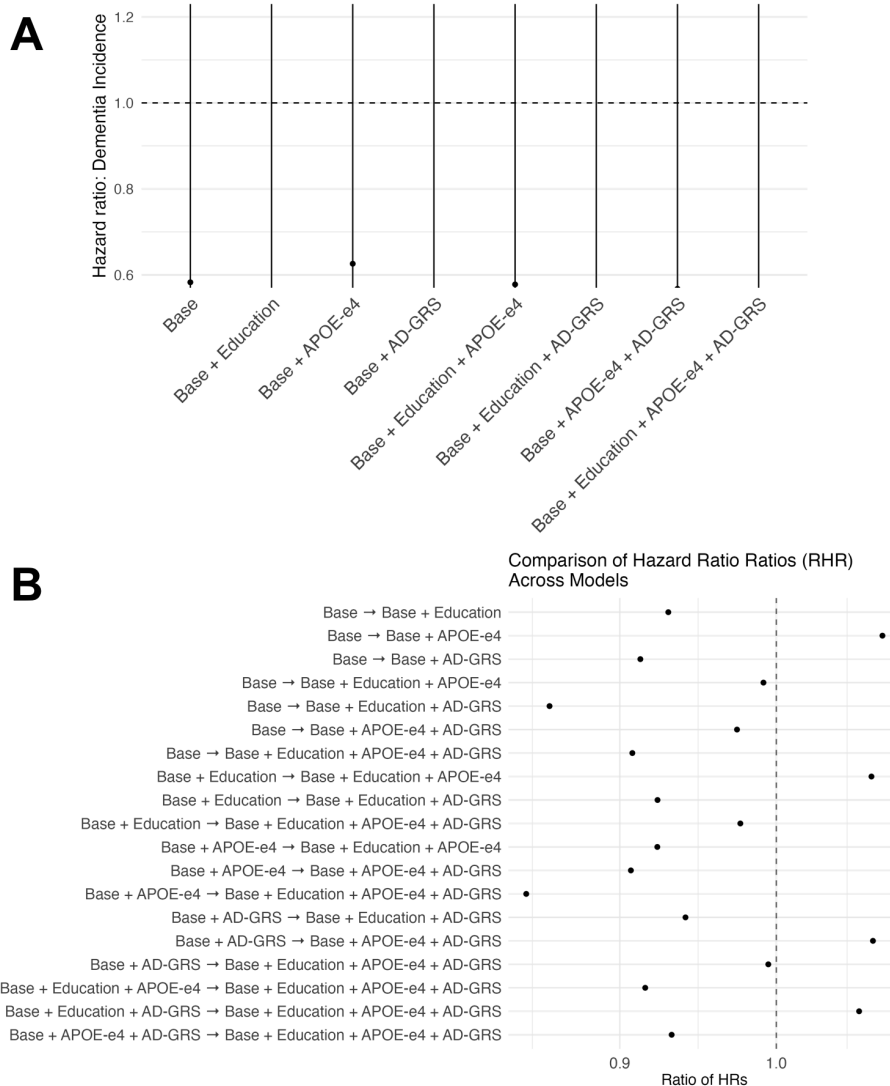

(A) Hazard ratios for dementia incidence comparing GLP-1 users with comparator therapies across progressively adjusted Cox proportional hazards models, including all combinations of confounders that are typically not adjusted for: education, APOE-ε4 genotype, and AD-GRS. (B) Ratios of hazard ratios (RHRs) comparing effect estimates between models, quantifying the relative change in the GLP-1-dementia association after additional covariate adjustment with typically unadjusted-for confounders. Points indicate point estimates; the vertical dashed line denotes no change (RHR = 1). Due to the small sample size, we did not perform bootstrap to obtain confidence intervals.

eFigure 10. Association between GLP-1 RA initiation versus SGLT2 inhibitor and dementia across nested models in individuals initiating second line therapy and with mild cognitive impairment.

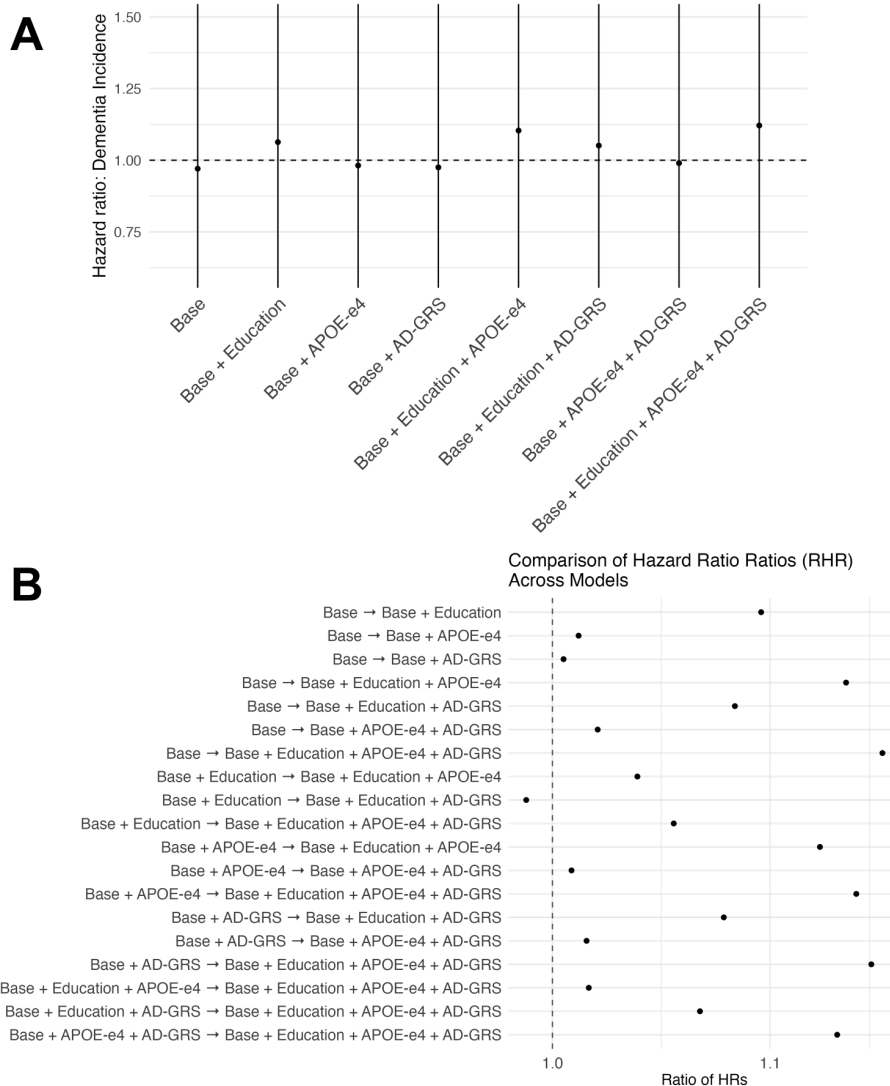

(A) Hazard ratios for dementia incidence comparing GLP-1 users with SGLT2 inhibitor use across progressively adjusted Cox proportional hazards models, including all combinations of confounders that are typically not adjusted for: education, APOE-ε4 genotype, and AD-GRS. (B) Ratios of hazard ratios (RHRs) comparing effect estimates between models, quantifying the relative change in the GLP-1-dementia association after additional covariate adjustment with typically unadjusted-for confounders. Points indicate point estimates; the vertical dashed line denotes no change (RHR = 1). Due to the small sample size, we did not perform bootstrap to obtain confidence intervals.
